# The statistical analysis of daily data associated with different parameters of the New Coronavirus COVID-19 pandemic in Georgia and their monthly interval prediction from January 1, 2022 to March 31, 2022

**DOI:** 10.1101/2022.04.19.22274044

**Authors:** Avtandil G. Amiranashvili, Ketevan R. Khazaradze, Nino D. Japaridze

## Abstract

In this work results of the next statistical analysis of the daily data associated with New Coronavirus COVID-19 infection of confirmed (C), recovered (R), deaths (D) and infection rate (I) cases of the population of Georgia in the period from January 01, 2022 to March 31, 2022 are presented. It also presents the results of the analysis of monthly forecasting of the values of C, D and I. As earlier, the information was regularly sent to the National Center for Disease Control & Public Health of Georgia and posted on the Facebook page https://www.facebook.com/Avtandil1948/.

The analysis of data is carried out with the use of the standard statistical analysis methods of random events and methods of mathematical statistics for the non-accidental time-series of observations. In particular, the following results were obtained.

Georgia’s ranking in the world for Covid-19 monthly mean values of infection and deaths cases in investigation period (per 1 million population) was determined. Among 157 countries with population ≥ 1 million inhabitants in February 2022 Georgia was in the 4 place on new infection cases, in the 2 place on death. Georgia took the best place in terms of confirmed cases of diseases (thirty fifth) and in mortality (tenth) - in March.

A comparison between the daily mortality from Covid-19 in Georgia from January 01, 2022 to March 31, 2022 with the average daily mortality rate in 2015-2019 shows, that the largest share value of D from mean death in 2015-2019 was 43.1 % (January 04, 2022), the smallest 2.12 % (March 23, 2022).

As in previous works [10–13] the statistical analysis of the daily and decade data associated with coronavirus COVID-19 pandemic of confirmed, recovered, deaths cases and infection rate of the population of Georgia are carried out. Maximum daily values of investigation parameters are following: C = 26320 (February 2, 2022), R = 48486 (February 12, 2022), D = 67 (January 4,2022), I = 41.58 % (February 14, 2022). Maximum mean decade values of investigation parameters are following: C = 22214 (1 Decade of February 2022), R = 23408 (2 Decade of February 2022), D = 45 (2 Decade of February 2022), I = 32.12% (1 Decade of February 2022).

It was found that as in spring, summer and from September to December 2021 [10,11,13], in investigation period of time the regression equations for the time variability of the daily values of C, R, D and I have the form of a tenth order polynomial.

Mean values of speed of change of confirmed - V(C), recovered - V(R), deaths - V(D) and infection rate V(I) coronavirus-related cases in different decades of months for the indicated period of time were determined. Maximum mean decade values of investigation parameters are following: V(C) = +1079 cases/day (3 Decade of January 2022), V(R) = +1139 cases/day (1 Decade of February 2022), V(D) = +0.8 cases/day (1 Decade of February 2022), V(I) = + 1.16 %/ day (3 decades of January 2022).

Cross-correlations analysis between confirmed COVID-19 cases with recovered and deaths cases shows, that from January 1, 2022 to March 31, 2022 the maximum effect of recovery is observed on 3-6 days after infection (CR=0.83-0.84), and deaths - after 2 and 4 days (CR=0.60). The impact of the omicron variant of the coronavirus on people (recovery, mortality) could be up to19 and 16 days respectively.

Comparison of daily real and calculated monthly predictions data of C, D and I in Georgia are carried out. It was found that in investigation period of time daily and mean monthly real values of C, D and I mainly fall into the 67% - 99.99% confidence interval of these predicted values. Exception - predicted values of I for January 2022 (alarming deterioration, violation of the stability of a time-series of observations).

Traditionally, the comparison of data about C and D in Georgia (GEO) with similar data in Armenia (ARM), Azerbaijan (AZE), Russia (RUS), Turkey (TUR) and in the World (WRL) is also carried out.

## 1. Introduction

More of two years have passed since the outbreak of the new coronavirus (COVID-19) in China, which was recognized on March 11, 2020 as a pandemic due to its rapid spread in the World [1]. During this period of time, despite the measures taken (including vaccination), several strains of this virus appeared (the last one is omicron). The overall level of morbidity and mortality in many countries of the world remains quite high. Scientists and specialists from various disciplines from all over the world continue intensive research on this unprecedented phenomenon (including in Georgia [2–13]), providing all possible assistance to epidemiologists.

In particular, in our work [13], it was noted works on statistical analysis, forecasting, forecasting systematization, spatial-temporary modeling of the spread of the new coronavirus etc. was actively continuing in 2021. Similar work is also continued in 2022 [14–17].

This work is a continuation of the researches [7–13].

In this work results of a statistical analysis of the daily data associated with New Coronavirus COVID-19 infection of confirmed (C), recovered (R), deaths (D) and infection rate (I) cases of the population of Georgia in the period from January 01, 2022 to March 31, 2022 are presented. It also presents the results of the analysis of monthly forecasting of the values of C, D and I. The information was regularly sent to the National Center for Disease Control & Public Health of Georgia and posted on the Facebook page https://www.facebook.com/Avtandil1948/.

The comparison of data about C and D in Georgia with similar data in Armenia, Azerbaijan, Russia, Turkey and in the world is also carried out.

We used standard methods of statistical analysis of random events and methods of mathematical statistics for non-random time series of observations [7–13, 18–20].

## 2. Study areas, material and methods

The study area: Georgia. Data of John Hopkins COVID-19 Time Series Historical Data (with US State and County data) [https://www.soothsawyer.com/john-hopkins-time-series-data-with-us-state-and-county-city-detail-historical/; https://data.humdata.org/dataset/total-covid-19-tests-performed-by-country] and https://stopcov.ge about daily values of confirmed, recovered, deaths and infection rate coronavirus-related cases, from January 01, 2022 to March 31, 2022 are used. The work also used data of National Statistics Office of Georgia (Geostat) on the average monthly total mortality in Georgia in January-December 2015-2019 [https://www.geostat.ge/en/]. In the proposed work the analysis of data is carried out with the use of the standard statistical analysis methods of random events and methods of mathematical statistics for the non-accidental time-series of observations [7–13, 18–20].

The following designations will be used below: Mean – average values; Min – minimal values; Max - maximal values; Range – Max-Min; St Dev - standard deviation; σ_m_-standard error; C_V_ = 100·St Dev/Mean – coefficient of variation, %; R^2^ – coefficient of determination; r – coefficient of linear correlation; CR – coefficient of cross correlation; Lag = 1, 2…60 Day; K_DW_ – Durbin-Watson statistic; Calc

– calculated data; Real - measured data; Dec1, Dec2, Dec3 – numbers of the month decades; α - the level of significance; C, R, D - daily values of confirmed, recovered and deaths coronavirus-related cases; V(C), V(R) and V(D) - daily values of speed of change of confirmed, recovered and deaths coronavirus-related cases (cases/day); I - daily values of infection rate (or positive rate) coronavirus-related cases (100· C/number of coronavirus tests performed); V(I) - daily values of speed of change of I; DC – deaths coefficient, % = (100· D/C). Official data on number of coronavirus tests performed are published from December 05, 2020 [https://stopcov.ge].

The statistical programs Data Fit 7, Mesosaur and Excel 16 were used for calculations.

The curve of trend is equation of the regression of the connection of the investigated parameter with the time at the significant value of the determination coefficient and such values of K_DW_, where the residual values are accidental. If the residual values are not accidental the connection of the investigated parameter with the time we will consider as simply regression.

The calculation of the interval prognostic values of C, D and I taking into account the periodicity in the time-series of observations was carried out using Excel 16 (the calculate methodology was description in [8]).

The duration and periodicity of time series of observations to calculating monthly forecasts of C, D and I values were as follows.

C values: for January-123 days, for February - 105 days, for March - 126 days. Periodicity – 7 days.

D values: for January-123 days, periodicity - 13 days; for February-110 days, periodicity - 13 days; for March–126 days, periodicity - 42 days

I values: for January - 123 days, periodicity – 7 days; for February - 105 days, periodicity – 6 days; for March – 123 days, periodicity – 6 days.

67%…99.99%_Low - 67%…99.99% lower level of confidence interval of prediction values of C, D and I; 67%…99.99%_Upp - 67%…99.99% upper level of confidence interval of prediction values of C, D and I.

In the Table 1 [9] the scale of comparing real data with the predicted ones and assessing the stability of the time series of observations in the forecast period in relation to the pre-predicted one (period for prediction calculating) is presented.

**Table 1.** The statistical characteristics of Covid-19 confirmed cases to 1 million populations in Georgia, neighboring countries (Armenia, Azerbaijan, Russia, Turkey) and World from January 1, 2022 to March 31, 2022 (r_min_ = ± 0.21, α = 0.05).

| Parameter | GEO | ARM | AZE | RUS | TUR | WRL |
| --- | --- | --- | --- | --- | --- | --- |
| Mean | 2115 | 290 | 190 | 553 | 703 | 281 |
| Min | 61 | 0 | 0 | 104 | 132 | 116 |
| Max | 7059 | 1478 | 761 | 1386 | 1307 | 519 |
| Range | 6998 | 1478 | 761 | 1282 | 1175 | 403 |
| Median | 1363 | 91 | 66 | 413 | 775 | 248 |
| St Dev | 2060 | 384 | 235 | 420 | 354 | 98 |
| $\sigma_m$ | 218.3 | 40.7 | 24.9 | 44.5 | 37.6 | 10.4 |
| $C_v$ (%) | 97.4 | 132.3 | 123.5 | 75.9 | 50.4 | 35.0 |
| Correlation Matrix |  |  |  |  |  |  |
| GEO | 1 | 0.94 | 0.93 | 0.76 | 0.87 | 0.45 |
| ARM | 0.94 | 1 | 0.90 | 0.72 | 0.77 | 0.41 |
| AZE | 0.93 | 0.90 | 1 | 0.84 | 0.78 | 0.26 |
| RUS | 0.76 | 0.72 | 0.84 | 1 | 0.70 | -0.05 |
| TUR | 0.87 | 0.77 | 0.78 | 0.70 | 1 | 0.54 |
| WRL | 0.45 | 0.41 | 0.26 | -0.05 | 0.54 | 1 |

## 3. Results and Discussion

The results in the Fig. 1–24 and Table 1–9 are presented.

**Fig. 1.**
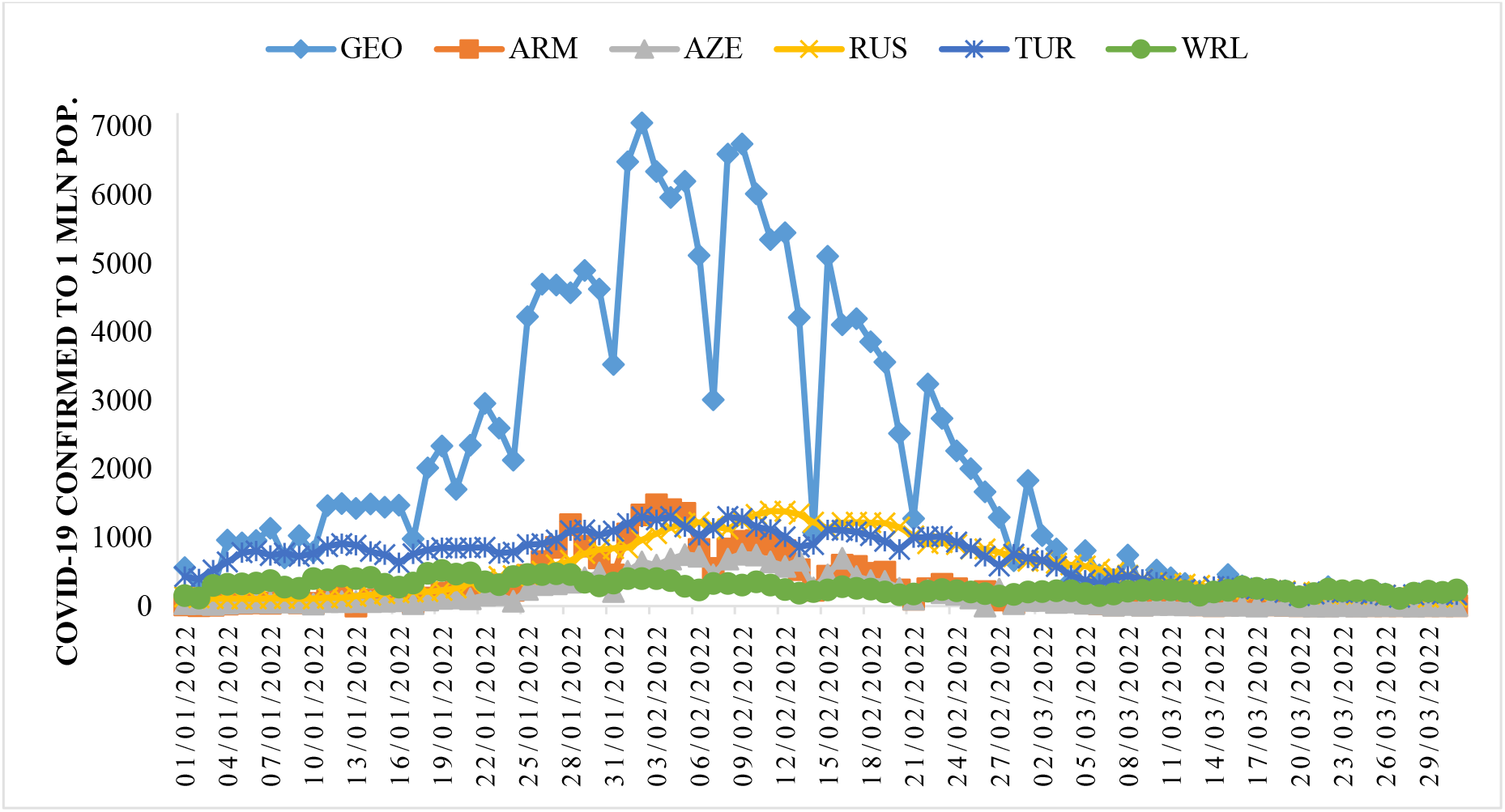
Time-series of Covid-19 confirmed cases to 1 million populations in Georgia, neighboring countries (Armenia, Azerbaijan, Russia, Turkey) and World from January 1, 2022 to March 31, 2022.

### 3.1 Comparison of time-series of Covid-19 confirmed and deaths cases in Georgia, its neighboring countries and World from January 1, 2022 to March 31, 2022

The time-series curves and statistical characteristics of Covid-19 confirmed and deaths cases (to 1 million populations) in Georgia, its neighboring countries and World from January 01, 2022 to March 31, 2022 in Fig. 1–2 and in Table 1–2 are presented.

**Fig. 2.**
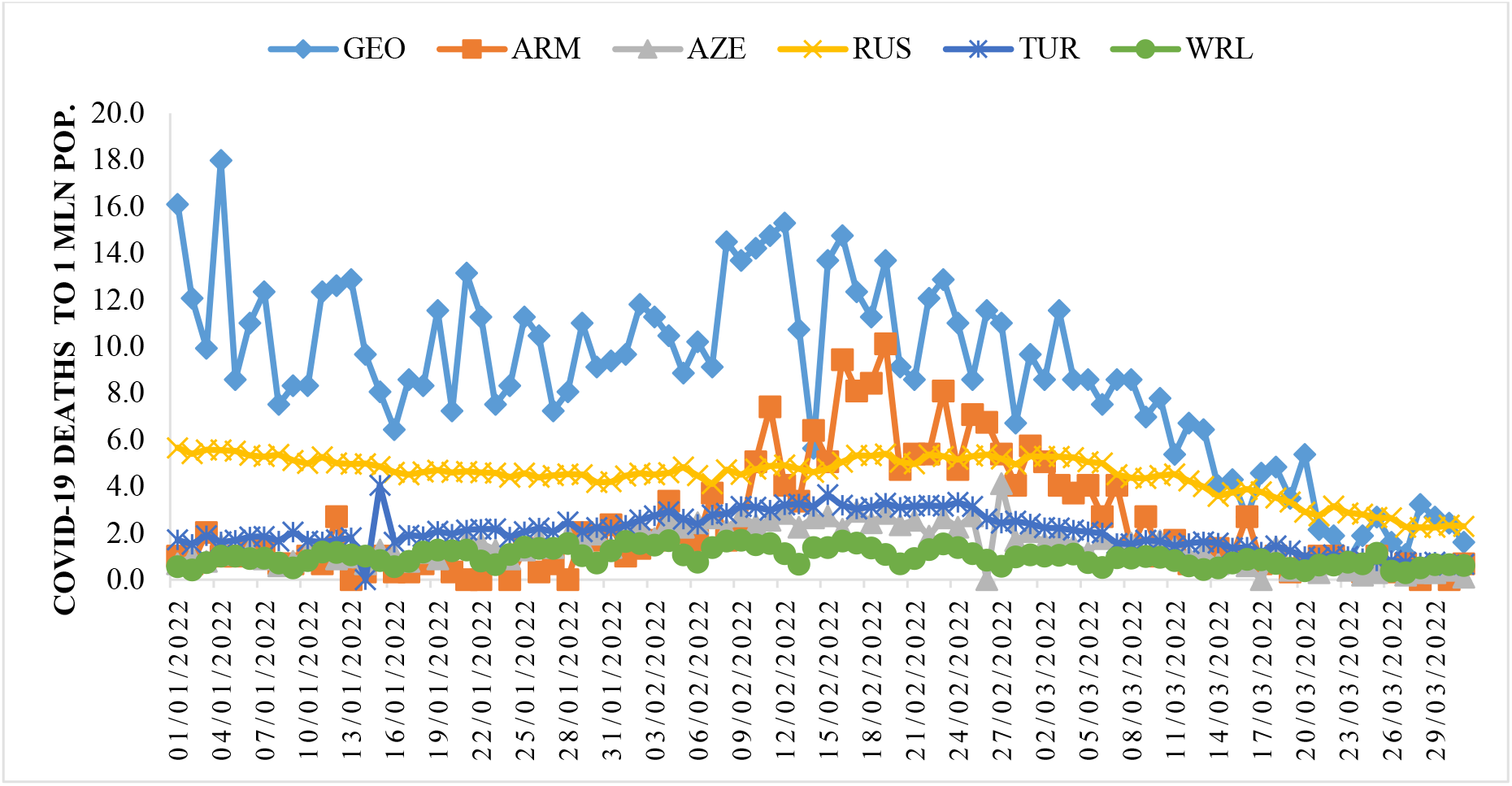
Time-series of deaths cases from Covid-19 to 1 million population in in Georgia, neighboring countries (Armenia, Azerbaijan, Russia, Turkey) and World from January 1, 2022 to March 31, 2022.

**Table 2.**
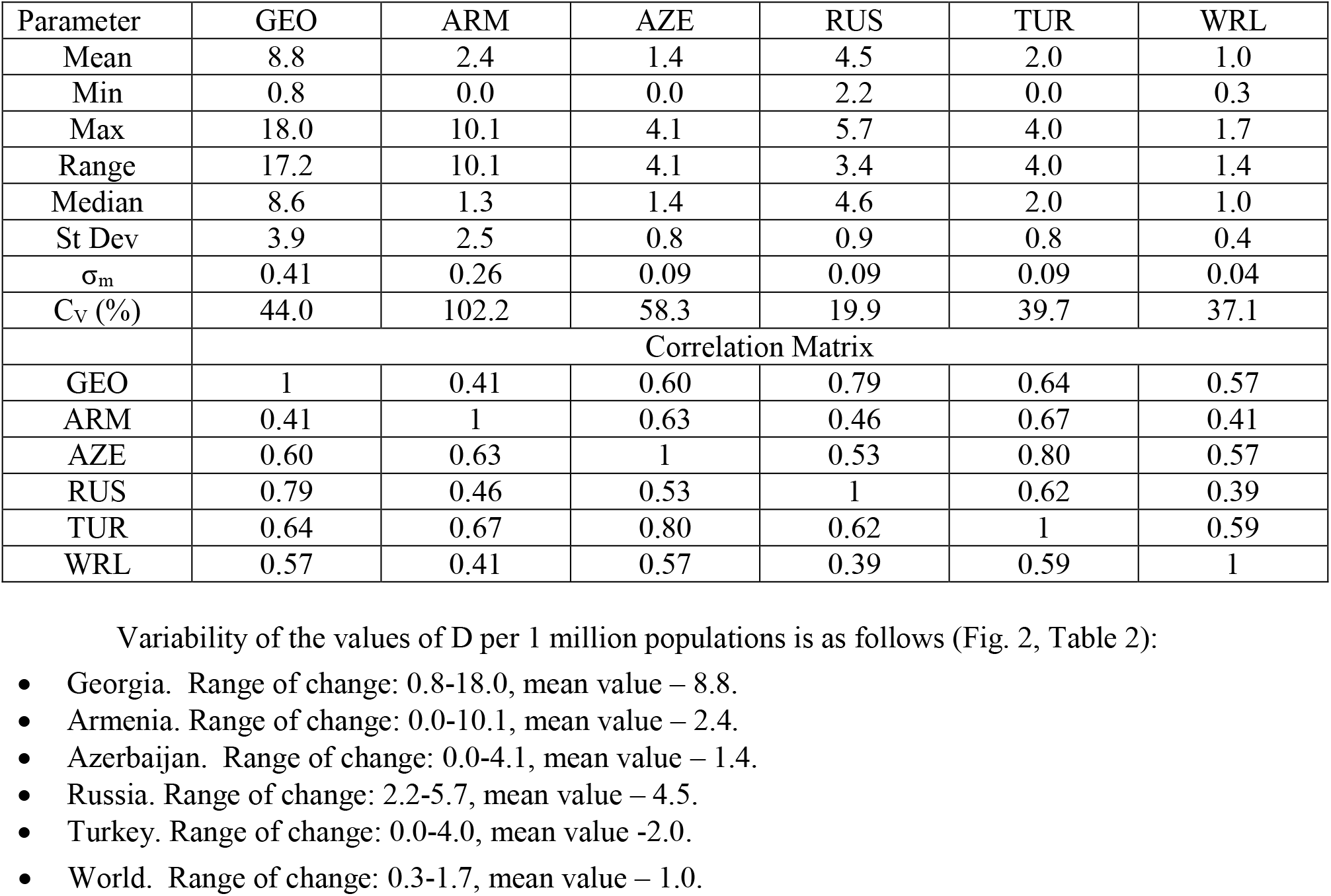
The statistical of deaths cases from Covid-19 to 1 million populations in Georgia, neighboring countries (Armenia, Azerbaijan, Russia, Turkey) and World from January 1, 2022 to March 31, 2022. (r_min_ = ± 0.21, α = 0.05).

Variability of the values of C per 1 million populations is as follows (Fig. 1, Table 1):

- Georgia. Range of change: 61-7059, mean value - 2115.
- Armenia. Range of change: 0-1478, mean value - 290.
- Azerbaijan. Range of change: 0-761, mean value - 190.
- Russia. Range of change: 104-1386, mean value - 553.
- Turkey. Range of change: 132-1307, mean value - 703.
- World. Range of change: 116-519, mean value - 281.

The largest variations in C values were observed in Armenia (C_V_=132.3%), the smallest - in Turkey (C_V_=50.4%).

Significant linear correlation (r_min_ = ± 0. 21, α = 0.05) between these countries on C value varies from 0.70 (pair Russia-Turkey) to 0.94 (pair Georgia-Armenia). Linear correlation between World and these countries is no significant only for pair World-Russia (r = −0.05).The degree of correlation [20] is as follows: very high correlation (0.9 ≤ R ≤ 1.0) - for pairs Georgia-Armenia, Georgia-Azerbaijan and Armenia-Azerbaijan; high correlation (0.7 ≤ R < 0.9)- for pairs Georgia-Russia, Georgia-Turkey, Armenia-Russia, Armenia-Turkey, Azerbaijan-Russia, Azerbaijan-Turkey and Russia-Turkey; moderate correlation (0.5 ≤ R < 0.7) – for pair World-Turkey; low correlation (0.3 ≤ R< 0.5) – for pairs World-Georgia and World-Armenia; negligible correlation (0 ≤ R < 0.3) – for pair World-Azerbaijan.

The largest variations in D values were observed in Armenia (C_V_=102.2 %), the smallest - in Russia (C_V_=19.9 %).

Significant linear correlation between these countries on D value varies from 0.41 (pair Georgia-Armenia) to 0.80 (pair Azerbaijan-Turkey). Linear correlation between World and these countries varies from 0.39 (pair World-Russia) to 0.59 (pair World-Turkey).

The degree of correlation [20] is as follows: high correlation - for pairs Georgia-Russia and Azerbaijan-Turkey; moderate correlation– for pairs Georgia-Azerbaijan, Georgia-Turkey, World-Georgia, Armenia-Azerbaijan, Armenia-Turkey, Azerbaijan-Russia, World-Azerbaijan, Russia-Turkey and World-Turkey; low correlation– for pairs Georgia-Armenia, Armenia-Russia, World-Armenia and World-Russia.

It should be noted that, in general, in the studied period of time, level of correlations shown in Tables 1 and 2 are higher than from September 1,2021 to December 31, 2021 [13].

In Table 3 the statistical characteristics of mean monthly values of C and D related to Covid-19 for 157 countries with population ≥ 1 million inhabitants from January to March 2022 (normed per 1 million population) is presented.

**Table 3.** The statistical characteristics of mean monthly values of confirmed cases and deaths cases related to Covid-19 for 157 countries with population ≥ 1 million inhabitant from January 2022 to March 2022 (normed on 1 mln pop.).

| Month | January |  | February |  | March |  |
| --- | --- | --- | --- | --- | --- | --- |
| Parameter | C | D | C | D | C | D |
| Mean | 679.5 | 1.60 | 652.8 | 2.04 | 416.9 | 1.31 |
| Min | 0.1 | 0 | 0.1 | 0 | 0 | 0 |
| Max | 5265.7 | 12.30 | 6280.4 | 11.72 | 6351.9 | 30.24 |
| Range | 5265.6 | 12.30 | 6280.2 | 11.72 | 6351.9 | 30.24 |
| St Dev | 1060.0 | 2.41 | 1089.4 | 2.72 | 903.5 | 2.80 |
| $\sigma_m$ | 84.9 | 0.19 | 87.2 | 0.22 | 72.3 | 0.22 |
| $C_V (\%)$ | 156 | 151 | 167 | 134 | 217 | 213 |

As follows from this Table range of change of mean monthly values of C for 157 countries varied from 0 (March) to 6352 (March). Average value of C for 157 countries varied from 417 (March) to 679.5 (January). Value of C_V_ changes from 156 % (January) to 217 % (March).

Range of change of mean monthly values of D for 157 countries varied from 0 (all months) to 30.24 (March). Average value of D for 157 countries varied from 1.31 (March) to 2.04 (February). Value of C_V_ changes from 134 % (February) to 213 % (March).

Between mean monthly values of Deaths and Confirmed cases related to Covid-19 for 157 countries with population ≥ 1 million inhabitant linear correlation and regression are observed (Fig. 3). As follows from Fig. 3 the highest growth rate of the monthly average values of D depending on C was observed in March, the smallest - in January (the corresponding values of the linear regression coefficients).

**Fig. 3.**
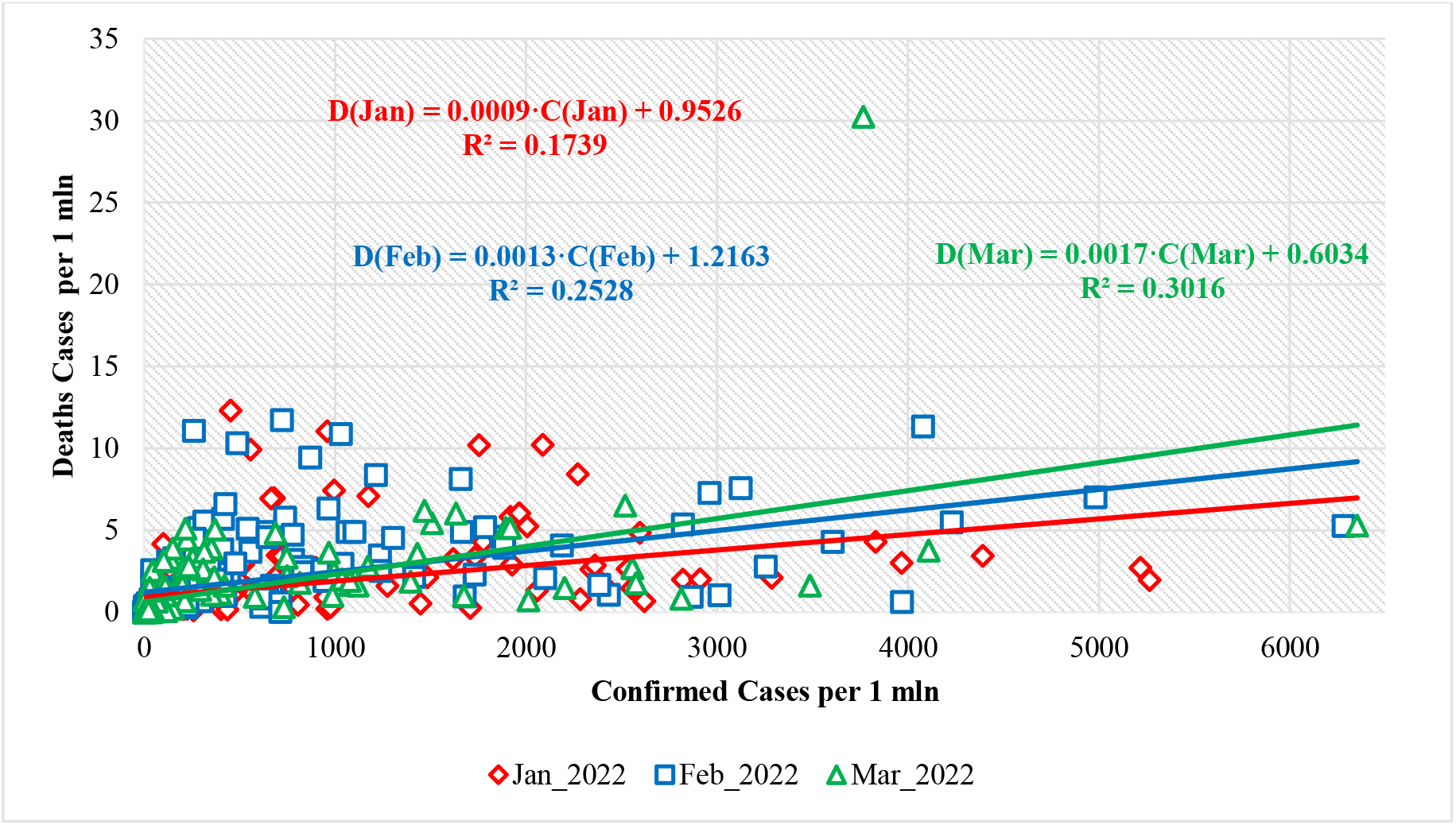
Linear correlation and regression between mean monthly values of deaths and confirmed cases related to Covid-19 for 157 countries with population ≥ 1 million inhabitants (normed on 1 mln pop.) from January 2022 to March 2022.

In Table 4 data about Covid-19 mean monthly values of infection (C) and deaths (D) cases from January to March 2022 (per 1 million population) and ranking of Georgia, Armenia, Azerbaijan, Russia and Turkey by these parameters (in brackets) among 157 countries with population ≥ 1 million inhabitant are presented. The corresponding values of the deaths coefficient (DC) are also given here.

**Table 4.** Covid-19 mean monthly values of infection (C) and deaths (D) cases from January to March 2022 (per 1 million population) and ranking of Georgia, Armenia, Azerbaijan, Russia, Turkey by these parameters (in brackets) among 157 countries with population ≥ 1 million inhabitant, and correspondent values of deaths coefficient (DC).

| Location | Georgia | Armenia | Azerbaijan | Russia | Turkey | World |
| --- | --- | --- | --- | --- | --- | --- |
| Parameter/Month | January |  |  |  |  |  |
| C (Rating C) | 2207 (18) | 249 (70) | 131 (82) | 298 (67) | 811 (41) | 366 |
| D ( Rating D) | 10.21 (3) | 0.88 (66) | 1.19 (62) | 4.86 (14) | 1.92 (47) | 0.97 |
| DC, % | 0.49 | 0.35 | 0.91 | 1.63 | 0.24 | 0.27 |
| Parameter/Month | February |  |  |  |  |  |
| C (Rating C) | 4079 (4) | 626 (50) | 444 (57) | 1099 (29) | 1037 (31) | 264 |
| D ( Rating D) | 11.33 (2) | 4.91 (23) | 2.39 (51) | 4.89 (26) | 2.95 (42) | 1.26 |
| DC, % | 0.28 | 0.78 | 0.54 | 0.44 | 0.28 | 0.48 |
| Parameter/Month | March |  |  |  |  |  |
| C (Rating C) | 370 (35) | 29 (77) | 20 (86) | 314 (39) | 292 (41) | 211 |
| D ( Rating D) | 5.04 (10) | 1.65 (39) | 0.87 (63) | 3.69 (16) | 1.36 (49) | 0.72 |
| DC, % | 1.36 | 5.66 | 4.39 | 1.17 | 0.47 | 0.34 |

In particular, as follows from this Table, mean monthly values of C for 5 country changes from 20 (Azerbaijan, March, 86 place between 157 country) to 4079 (Georgia, February, 4 place between 157 country).

Mean monthly values of D for 5 country changes from 0.87 (Azerbaijan, March, 63 place between 157 country) to 11.33 (Georgia, February, 2 place between 157 country).

On the whole (Table 4) in February 2022 Georgia was in the 4 place on new infection cases, and in in the 2 place on death. Georgia took the best place in terms of confirmed cases of diseases (thirty fifth) and in mortality (tenth) – in March.

Mean monthly values of DC for 5 country changes from 0.24% (Turkey, January) to 5.66 % (Armenia, March).

Note that the mean values of DC (%) from January to March 2022 are: Georgia – 0.41, Armenia – 0.83, Azerbaijan – 0.76, Russia – 0.81, Turkey – 0.29, World – 0.35 (according to data from Table 1 and 2).

The mean values of DC (%) from September to December 2021 were: Georgia – 1.67, Armenia – 3.05, Azerbaijan – 1.42, Russia – 3.50, Turkey – 0.83, World – 1.31 [13].

In summer 2021 the mean values of DC (%) were: Georgia – 1.28, Armenia – 2.08, Azerbaijan – 0.80, Russia – 3.36, Turkey – 0.80, World – 1.79 [11]. Thus, during the study period, compared to the previous two, the DC values in all five indicated countries and World has decreased.

### 3.2 Comparison of daily death from Covid-19 in Georgia with daily mean death in 2015-2019 from January 1, 2022 to March 31, 2022

In Fig. 4–7 data of the daily death from Covid-19 in Georgia from October 1, 2020 to March 31, 2022 in comparison with daily mean death in 2015-2019 are presented. The daily mean death in 2015-2019 in different months are: January - 155, February – 146, March - 141, April - 137, May – 131, June – 124, July – 117, August – 120, September – 112, October - 123, November - 135, December - 144 (Fig. 4, 6).

**Fig. 4.**
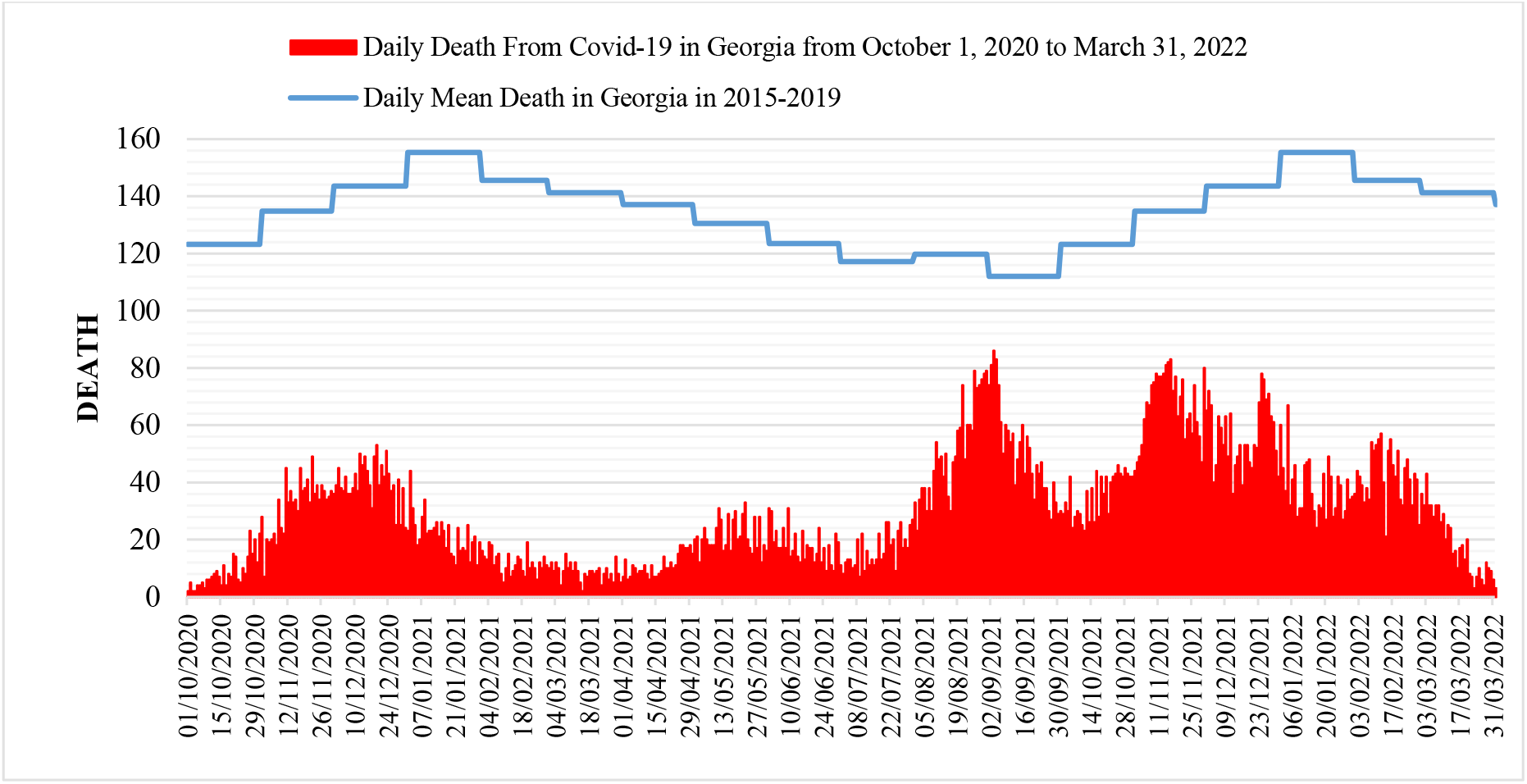
Daily death from Covid-19 in Georgia from October 1, 2020 to March 31, 2022 in comparison with daily mean death in 2015-2019.

The daily death from Covid-19 from January 1, 2022 to March 31, 2022 (Fig. 4) changes from 3 (March 23, 2022) to 67 (January 4, 2022). A comparison between the daily mortality from Covid-19 in Georgia from January 01, 2022 to March 31, 2022 with the average daily mortality rate in 2015-2019 shows, that the largest share value of D from mean death in 2015-2019 (Fig. 5) was 43.1 % (January 04, 2022), the smallest - 2.12 % (March 23, 2022).

**Fig. 5.**
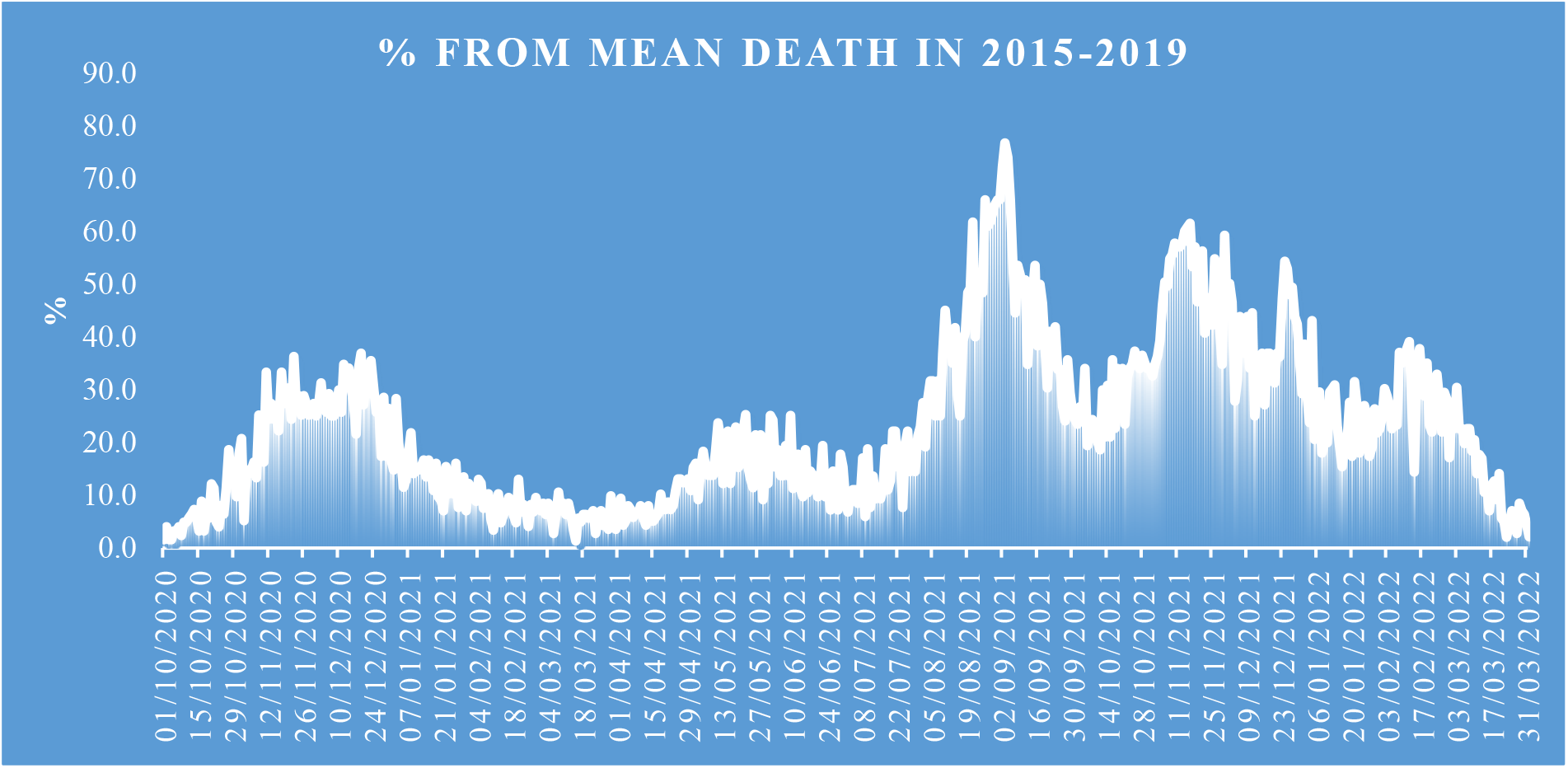
Share value of D from mean death in 2015-2019.

The most share of mean daily mortality from Covid-19 of mean daily mortality in 2015-2019 from October 2020 to March 2022 in November was observed – 49.7% (Fig. 7).

**Fig. 6.**
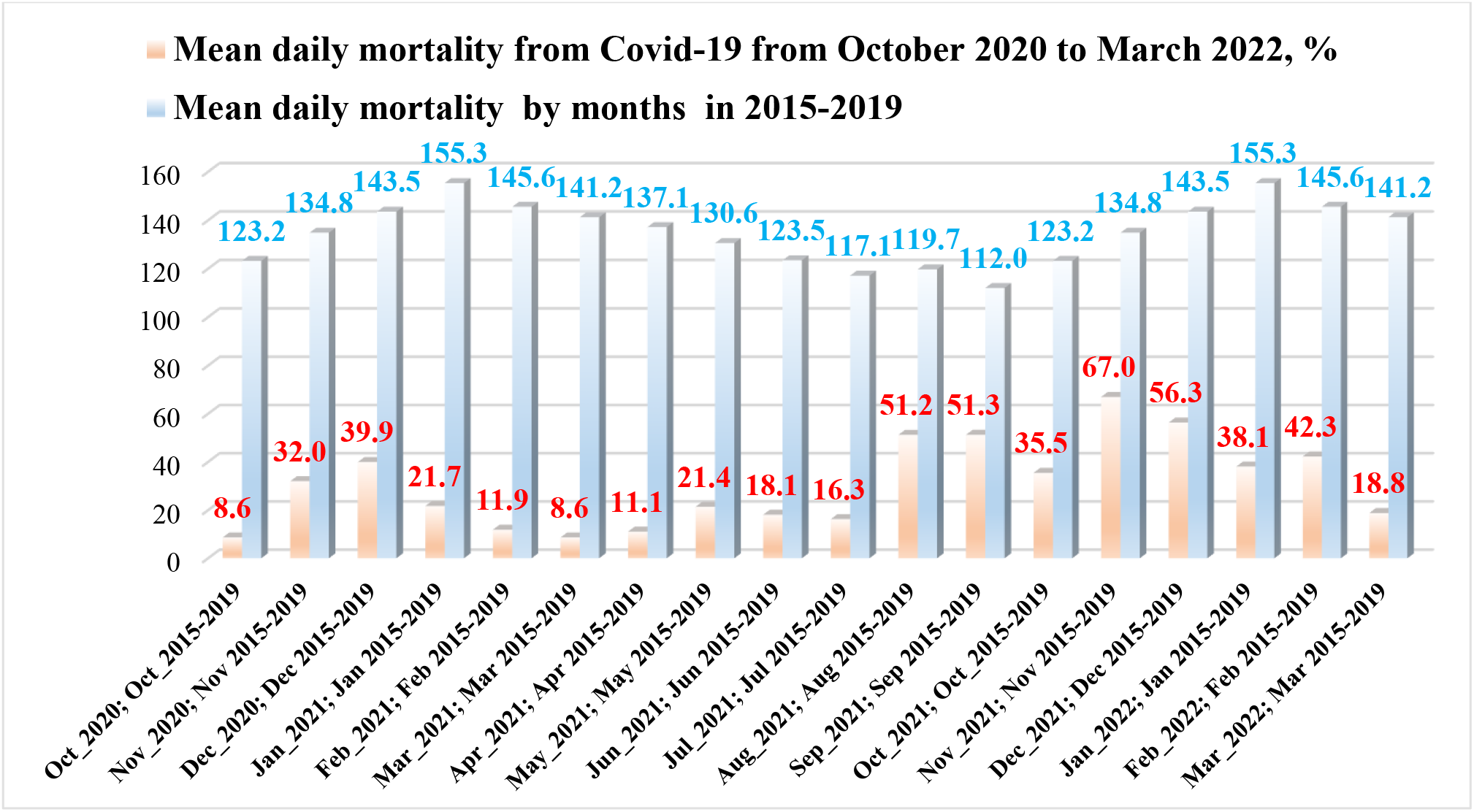
Mean daily mortality from Covid-19 from October 2020 March 2022, % and mean daily mortality from January to December in 2015-2019.

**Fig. 7.**
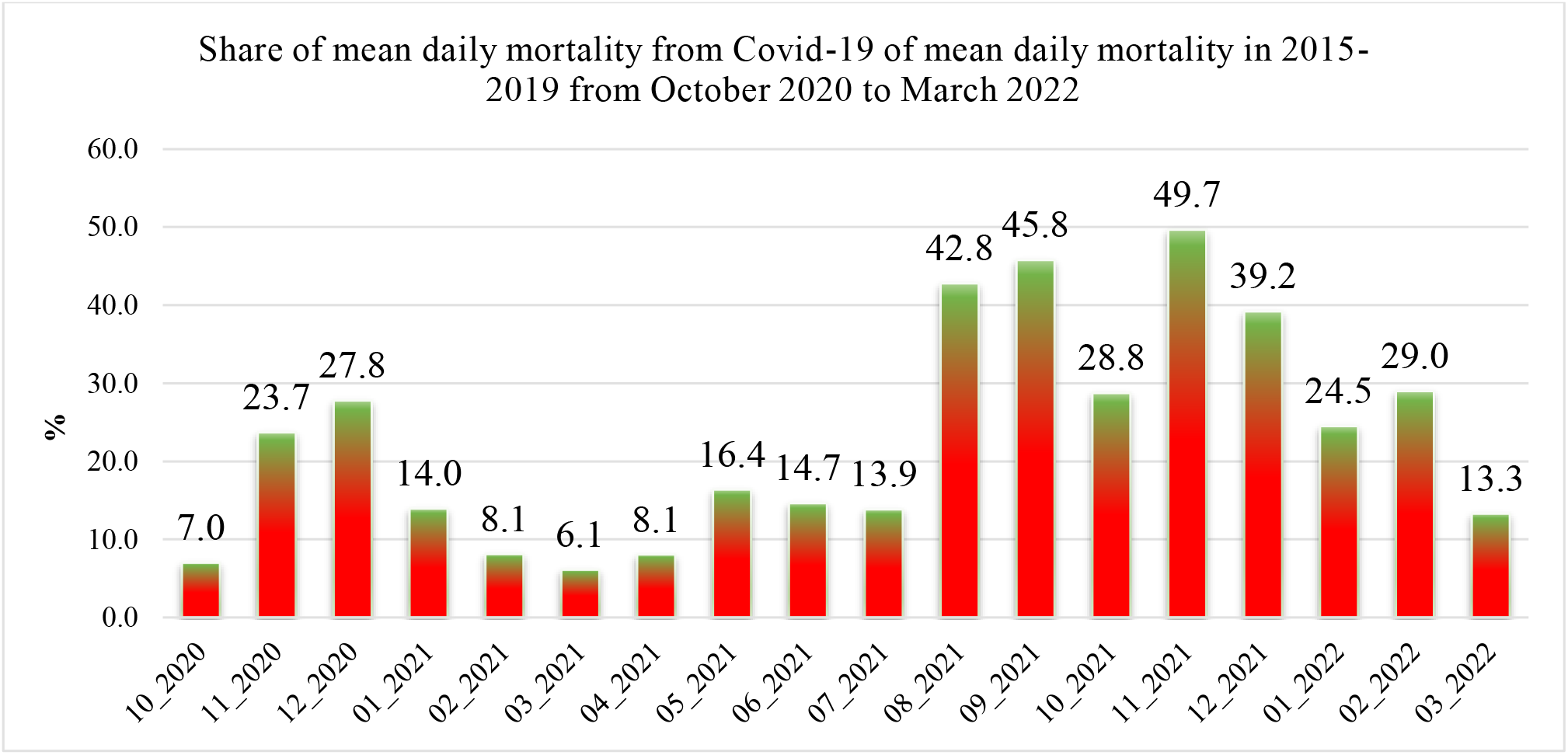
Share of mean daily mortality from Covid-19 of mean daily mortality in 2015-2019 from October 2020 to March 2021.

### 3.3 The statistical analysis of the daily and decade data associated with New Coronavirus COVID-19 pandemic from January to March 2022

Results of the statistical analysis of the daily and decade data associated with New Coronavirus COVID-19 pandemic in Georgia from January 1, 2022 to March 31, 2022 in Tables 5–7 and Fig. 8 – 20 are presented.

**Table 5.** Statistical characteristics of the daily data associated with coronavirus COVID-19 pandemic of confirmed, recovered, deaths cases and infection rate of the population of Georgia from 01.01.2021 to 31.03.2022.

| Parameter | Confirmed | Recovered | Deaths | Infection Rate, % |
| --- | --- | --- | --- | --- |
| Mean | 7887 | 8108 | 32.7 | 16.98 |
| Min | 229 (Mar 28) | 777 (Jan 9) | 3 (Mar 23) | 1.90 (Mar 31) |
| Max | 26320 (Feb 2) | 48486 (Feb 12) | 67 (Jan 4) | 41.58 (Feb 14) |
| Range | 26091 | 47709 | 64 | 39.67 |
| St Dev | 7680 | 7995 | 14.4 | 10.40 |
| $\sigma_m$ | 814 | 847 | 1.5 | 1.10 |
| $C_v$ (%) | 97.4 | 98.6 | 44.0 | 61.2 |

**Fig. 8.**
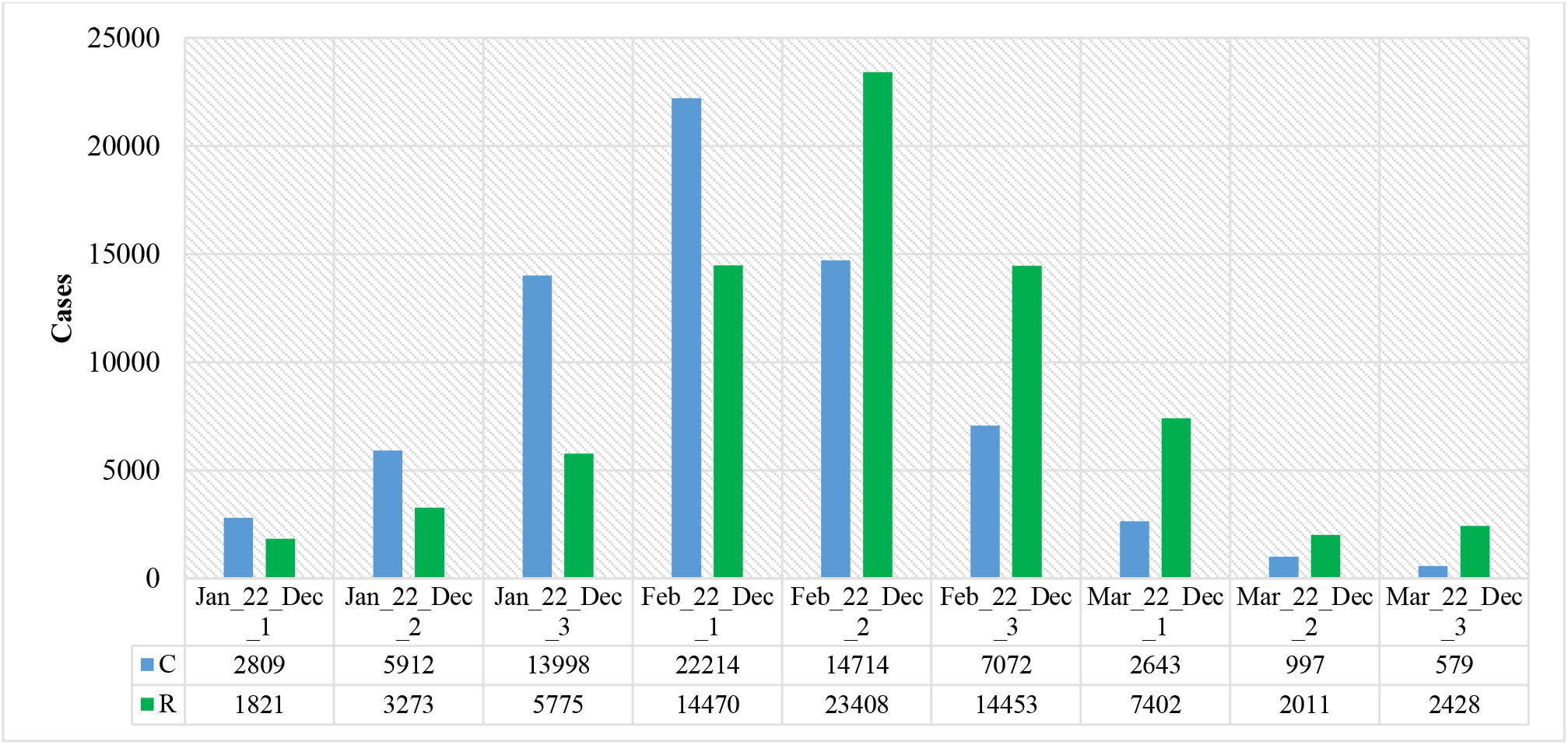
Mean values of confirmed and recovered coronavirus-related cases in different decades of months in Georgia from January 2022 to March 2022.

The mean and extreme values of the studied parameters are as follows (Table 5): C - mean - 7887, range: 229-26320; R - mean - 8108, range: 777 - 48486; D - mean - 32.7, range: 3-67; I (%) - mean – 16.98, range: 1.90 – 41.58. The range of variability for all studied parameters44.0% (D) ≤C_**V**_≤98.6% (R).

Mean decade values of confirmed and recovered coronavirus-related cases varies within the following limits (Fig. 8): C – from 579 (3 Decade of March 2022) to 22214 (1 Decade of February 2022); R – from 1821 (1 Decade of January 2022) to 23408 (2 Decade of February 2022).

Mean decade values of deaths coronavirus-related cases (Fig. 9) varies from 7 (3 Decade of March) to 45 (2 Decade of February).

**Fig. 9.**
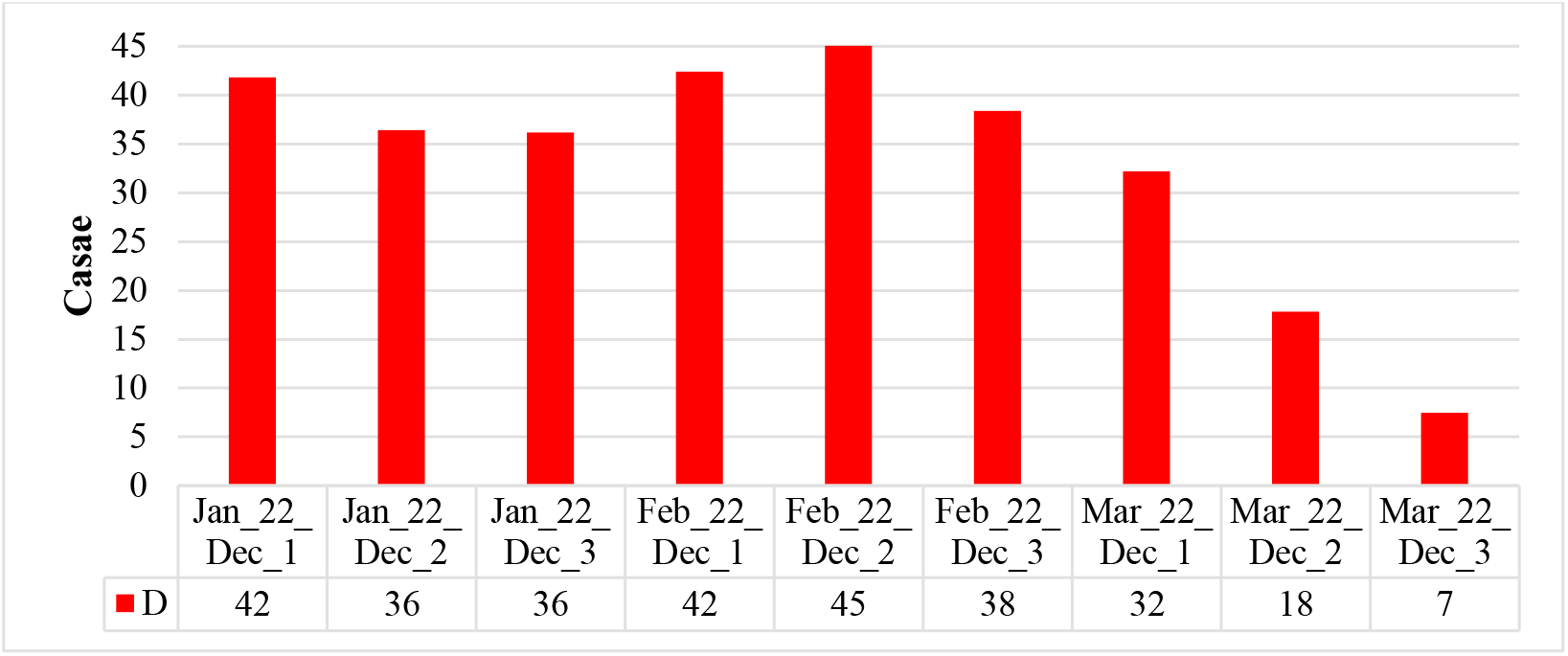
Mean values of deaths coronavirus-related cases in different decades of months in Georgia from January 2022 to March 2022.

Mean decade values of infection rate coronavirus-related cases (Fig. 10) varies from 3.51 % (3 Decade of March) to 32.12 % (1 Decade of February).

**Fig. 10.**
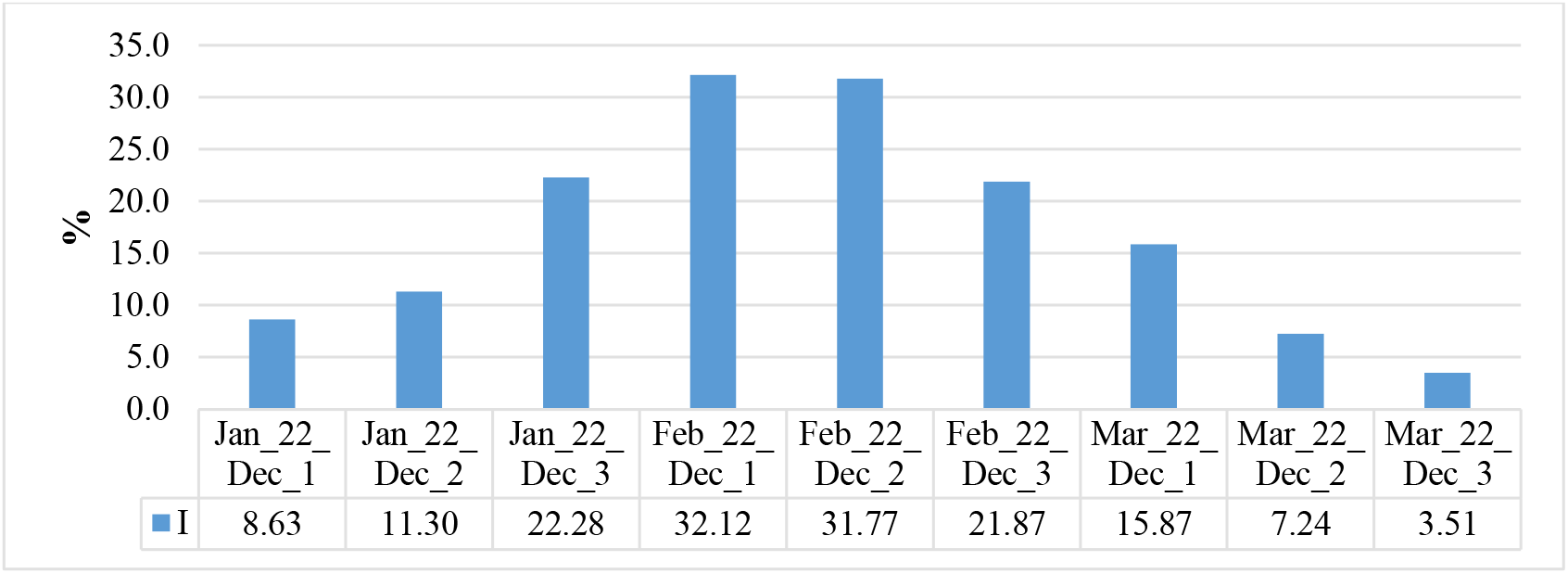
Mean values of infection rate coronavirus-related cases in different decades of months in Georgia from January 2022 to March 2022.

Time changeability of the daily values of C, R, D and I are satisfactorily described by the tenth order polynomial (Table 6, Fig. 11–13). For clarity, the data in Fig. 12 are presented in relative units (%) in relation to their average values.

**Table 6.** Form of the equations of the regression of the time changeability of the daily values of C, R and D from January 01, 2022 to March 31, 2022 in Georgia. The level of significance of R^2^ is not worse than 0.001.

| Parameter | Confirmed | Recovered | Deaths | Infection Rate |
| --- | --- | --- | --- | --- |
| Regression | Tenth Order Polynomial |  |  |  |
| $R^2$ | 0.894 | 0.784 | 0.753 | 0.972 |
| $K_{DW}$ | 1.93 | 1.83 | 2.02 | 1.63 |
| Autocorrelation of residual components | Absent |  |  |  |

**Fig. 11.**
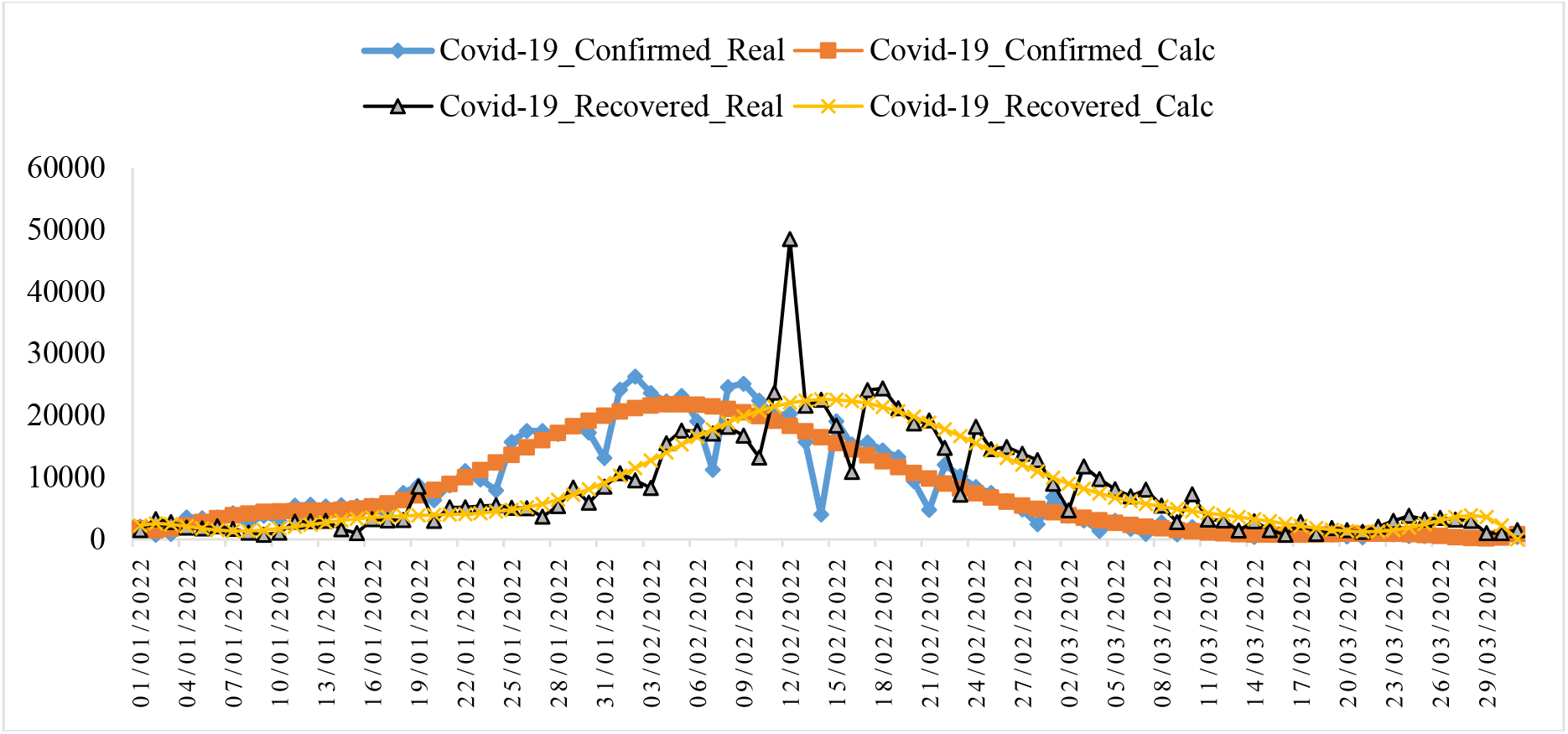
Changeability of the real and calculated daily values of coronavirus confirmed and recovered cases from January 01, 2022 to March 31, 2022 in Georgia.

**Fig. 12.**
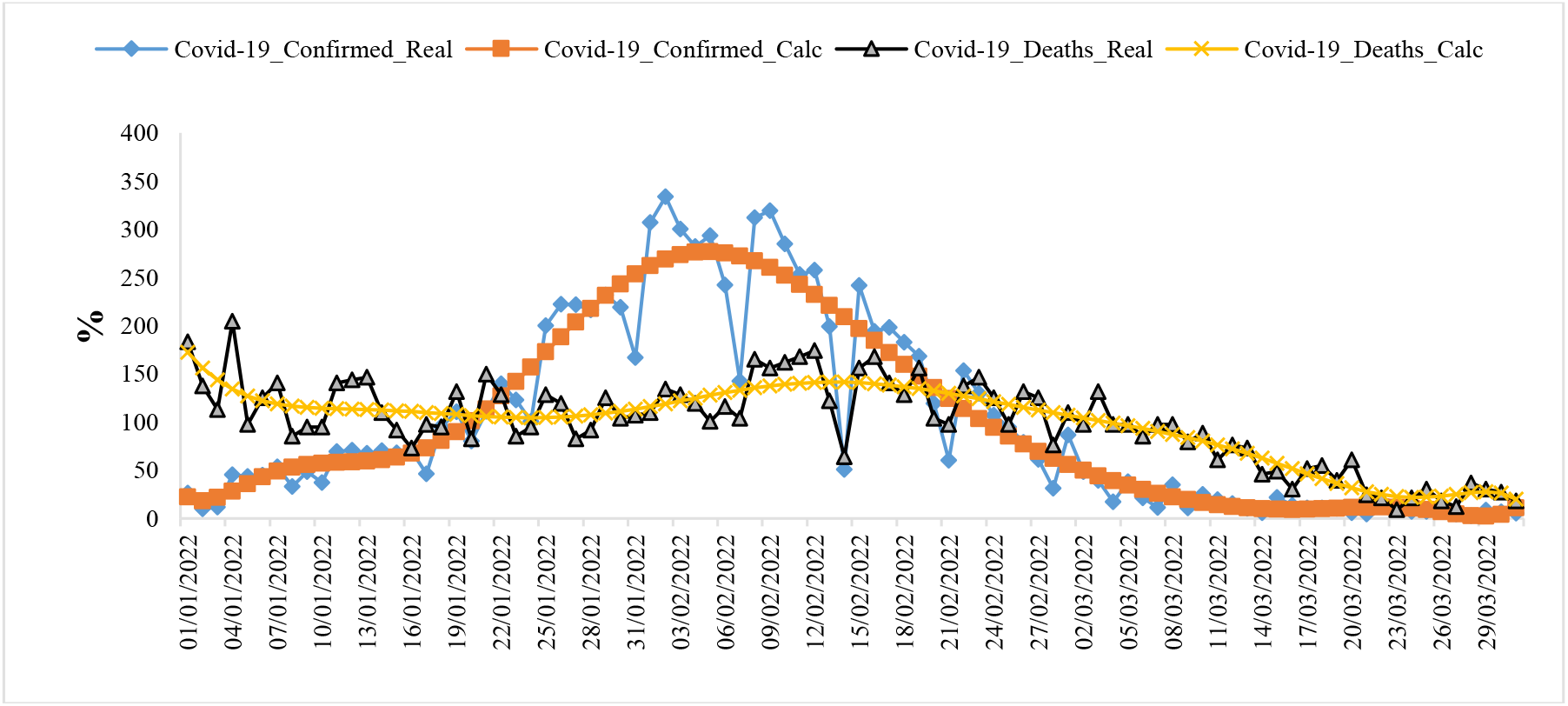
Changeability of the real and calculated daily values of coronavirus-related confirmed and deaths cases from January 01, 2022 to March 31, 2022 in Georgia (normed on mean values, %).

**Fig. 13.**
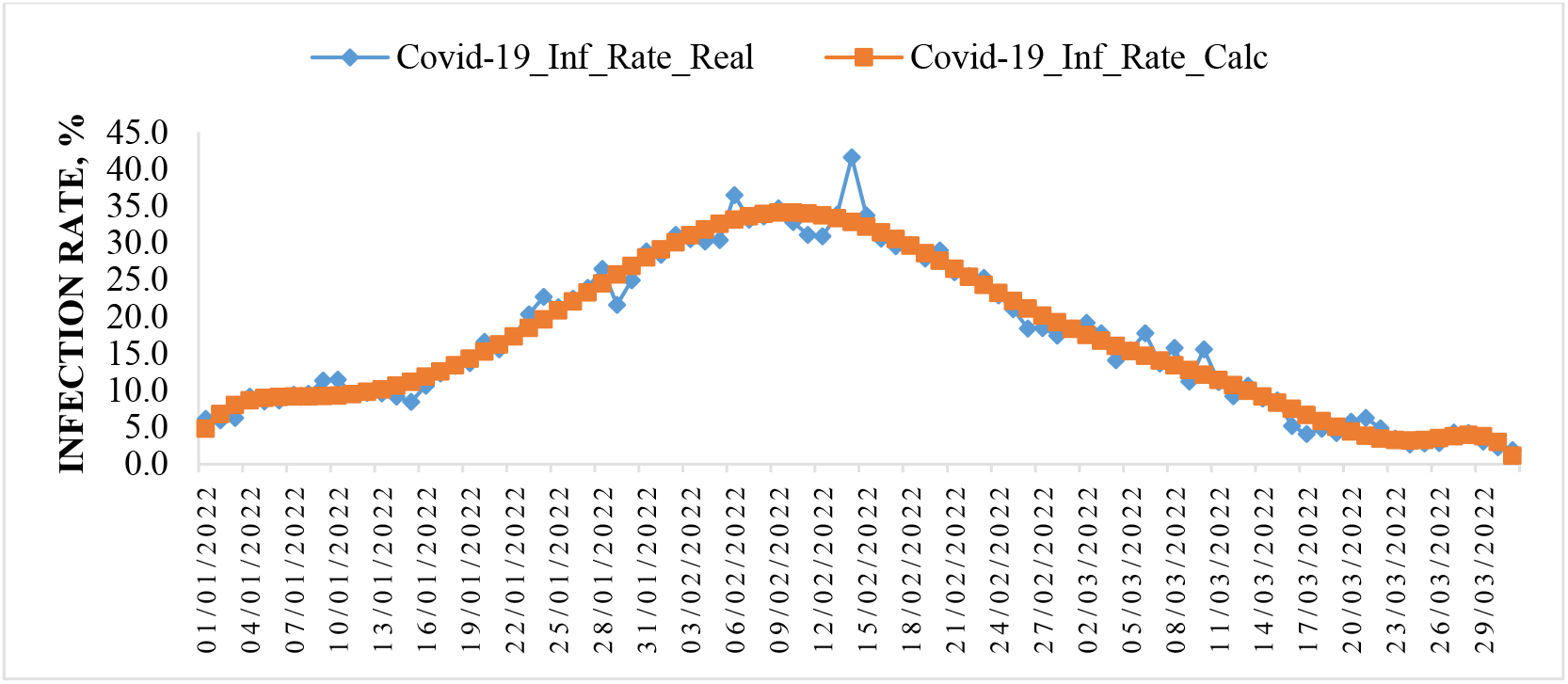
Changeability of the real and calculated daily values of coronavirus Infection Rate from January 01, 2022 to March 31, 2022 in Georgia.

Note that from Fig. 11 and 12, as in [8–13], clearly show the shift of the time series values of R and D in relation to C.

In Fig. 14–16 data about mean values of speed of change of confirmed, recovered, deaths coronavirus-related cases and infection rate in different decades of months from January to March 2022 are presented.

**Fig. 14.**
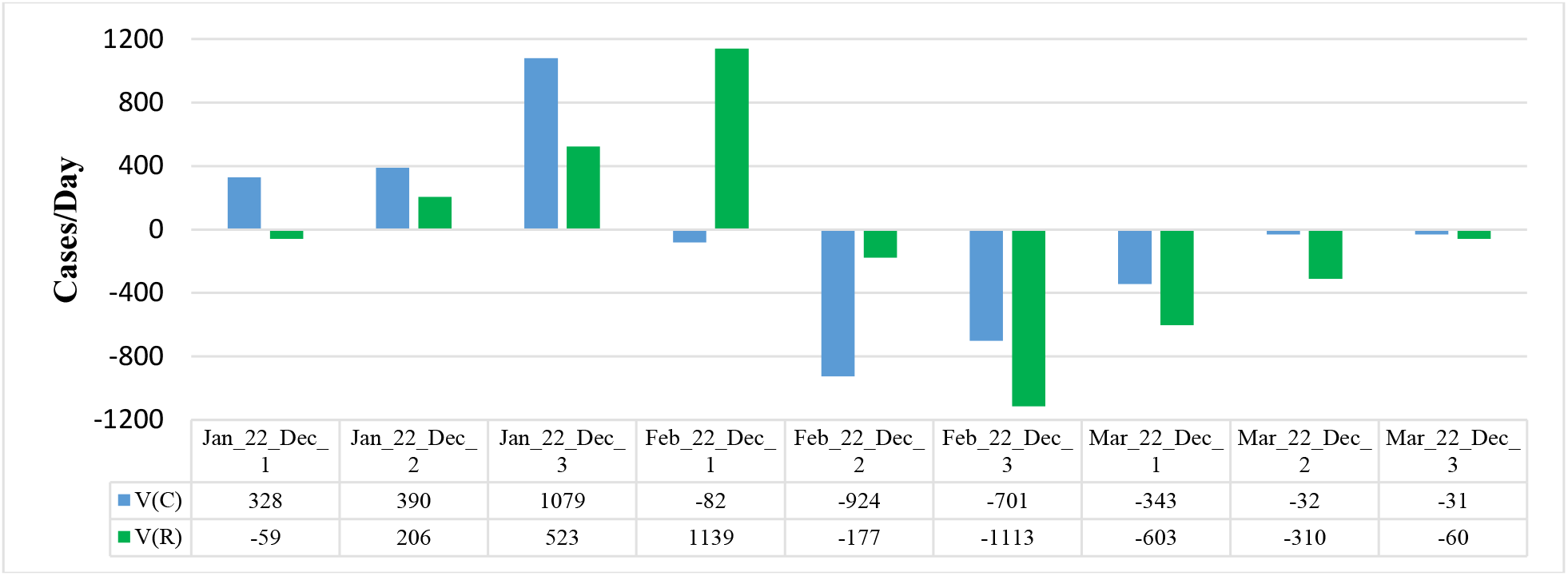
Mean values of speed of change of confirmed and recovered coronavirus-related cases in different decades of months in Georgia from January 2022 to March 2022.

Maximum mean decade values of investigation parameters are following: V(C) = +1079 cases/day (3 Decade of January), V(R) = +1139 cases/day (1 Decade of February), V(D) = +0.8 cases/day (1 Decade of February), V(I) = + 1.16 %/ day (3 Decade of January). Min mean decade values of investigation parameters are following: V(C) = −924 cases/day (2 Decade of February), V(R) = −1113 cases/day (3 Decade of February), V(D) = −1.8 cases/day (1 Decade of January), V(I) = −1.03 %/day (3 Decade of February).

Data about mean monthly values of C, R, D, I and its speed of change in from January to March 2022 in Table 7 are presented. As follows from this Table there was an increase in average monthly values of C, R, D and I from January to February, and further decrease to March.

**Table 7.** Mean monthly values of C, R, D, I and its speed of change in Georgia from January 2022 to March 2022.

| Parameter | C | R | D | I, % | V(C) | V(R) | V(D) | V(I) |
| --- | --- | --- | --- | --- | --- | --- | --- | --- |
| Jan_2022 | 7780 | 3692 | 38.1 | 14.34 | 623.9 | 242.6 | -0.5 | 0.76 |
| Feb_2022 | 15209 | 17657 | 42.3 | 29.07 | -559.6 | 25.7 | -0.1 | -0.35 |
| Mar_2022 | 1380 | 3898 | 18.8 | 8.70 | -135.5 | -324.4 | -0.9 | -0.56 |

The values of V(C), V(R), V(D) and V(I) changes as follows: V(C) – from −560 cases/day (February) to +624 cases/day (January); V(R) - from −324 cases/day (March) to +243 cases/day (January); V(D) - from −0.9 cases/day (March) to −0.1 cases/day (February) and V(I) - from −0.56 %/day (March) to +0.76 %/day (January).

In Fig. 17 data about connection of 14-day moving average of deaths cases due to COVID-19 in Georgia with 14-day moving average of infection rate from December 18, 2020 until March 31, 2022 are presented. As follows from Fig. 17, in general, with an increase of the infection rate is observed increase of deaths cases.

**Fig. 15.**
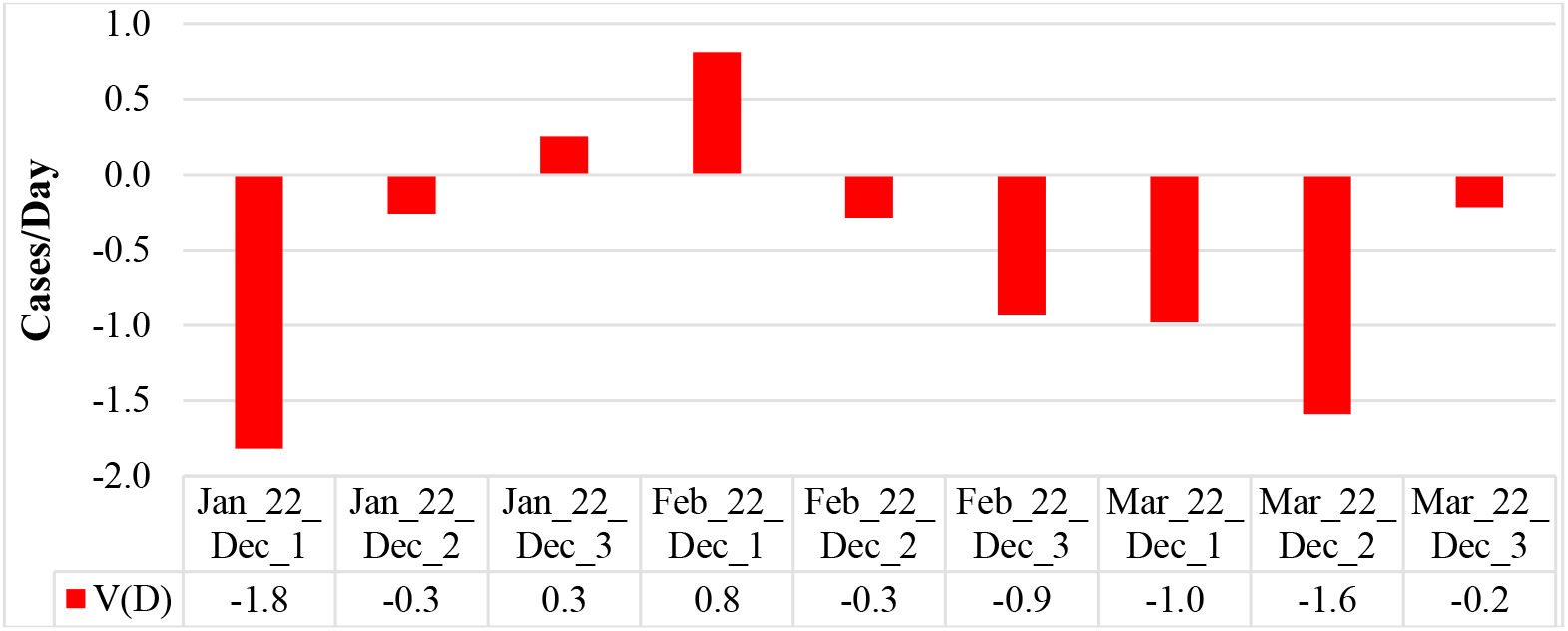
Mean values of speed of change of deaths coronavirus-related cases in different decades of months in Georgia from January 2022 to March 2022.

**Fig. 16.**
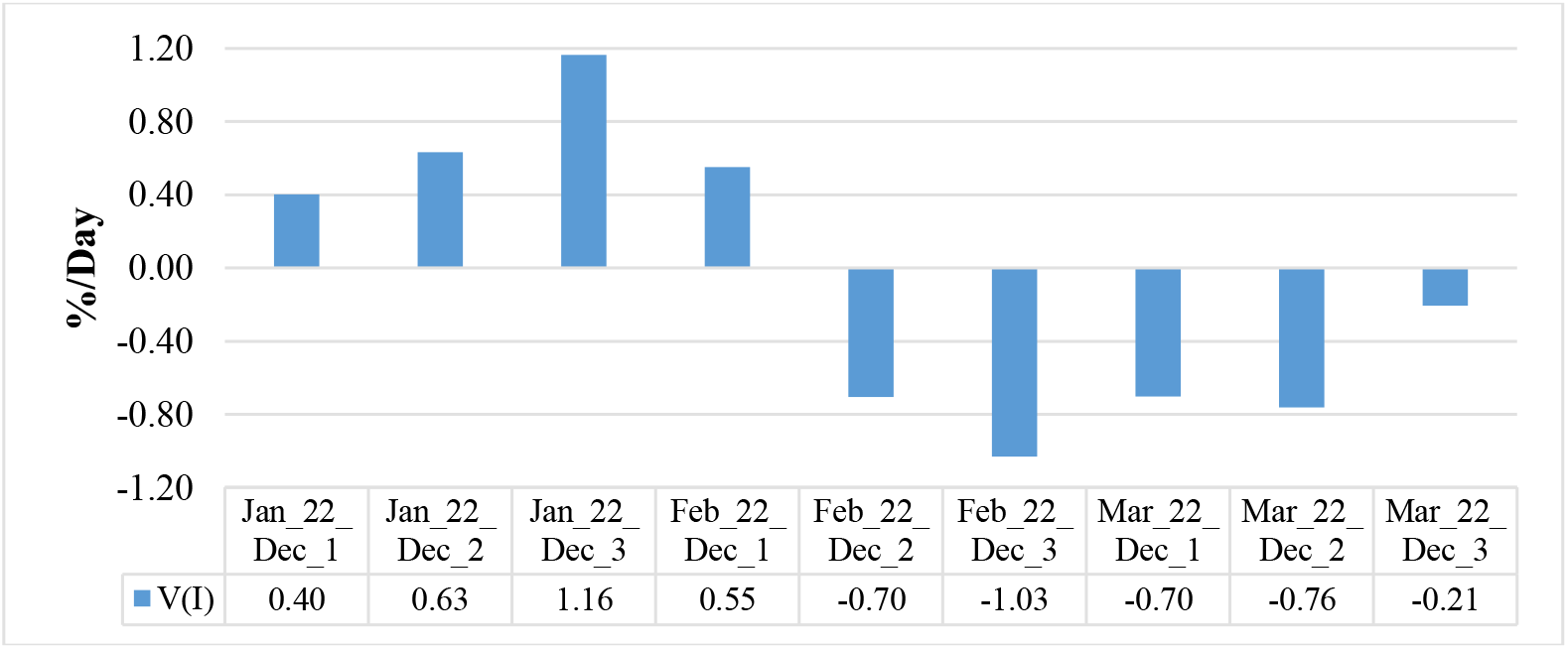
Mean values of speed of change of coronavirus infection rate in different decades of months in Georgia from January 2022 to March 2022.

**Fig. 17.**
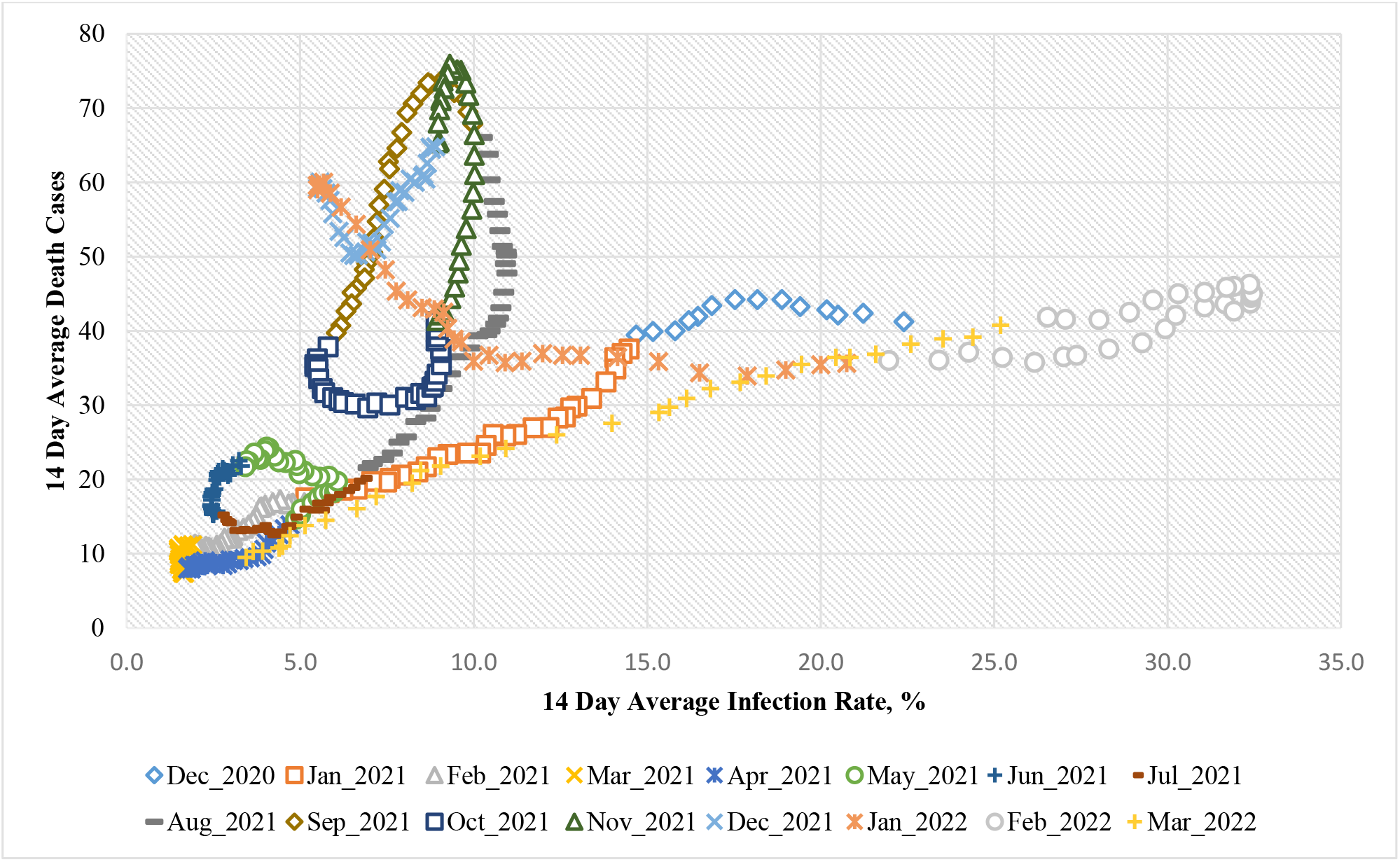
Connection of 14-day moving average of deaths cases due to COVID-19 in Georgia with 14-day moving average of infection rate from December 18, 2020 until March 31, 2022.

R=0.45 (low correlation [20]), α≈0.075

Using the data in Fig. 18, a linear regression graph between the monthly mean values of D and I is obtained (Fig. 19). As follows from Fig. 19, in general a low level of linear correlation between these parameters is observed. This Fig. also clearly demonstrates the anomaly high mortality from coronavirus in November 2021 with relatively low value of infection rate in comparison with February 2022, which reduces the level of correlation between D and I.

**Fig. 18.**
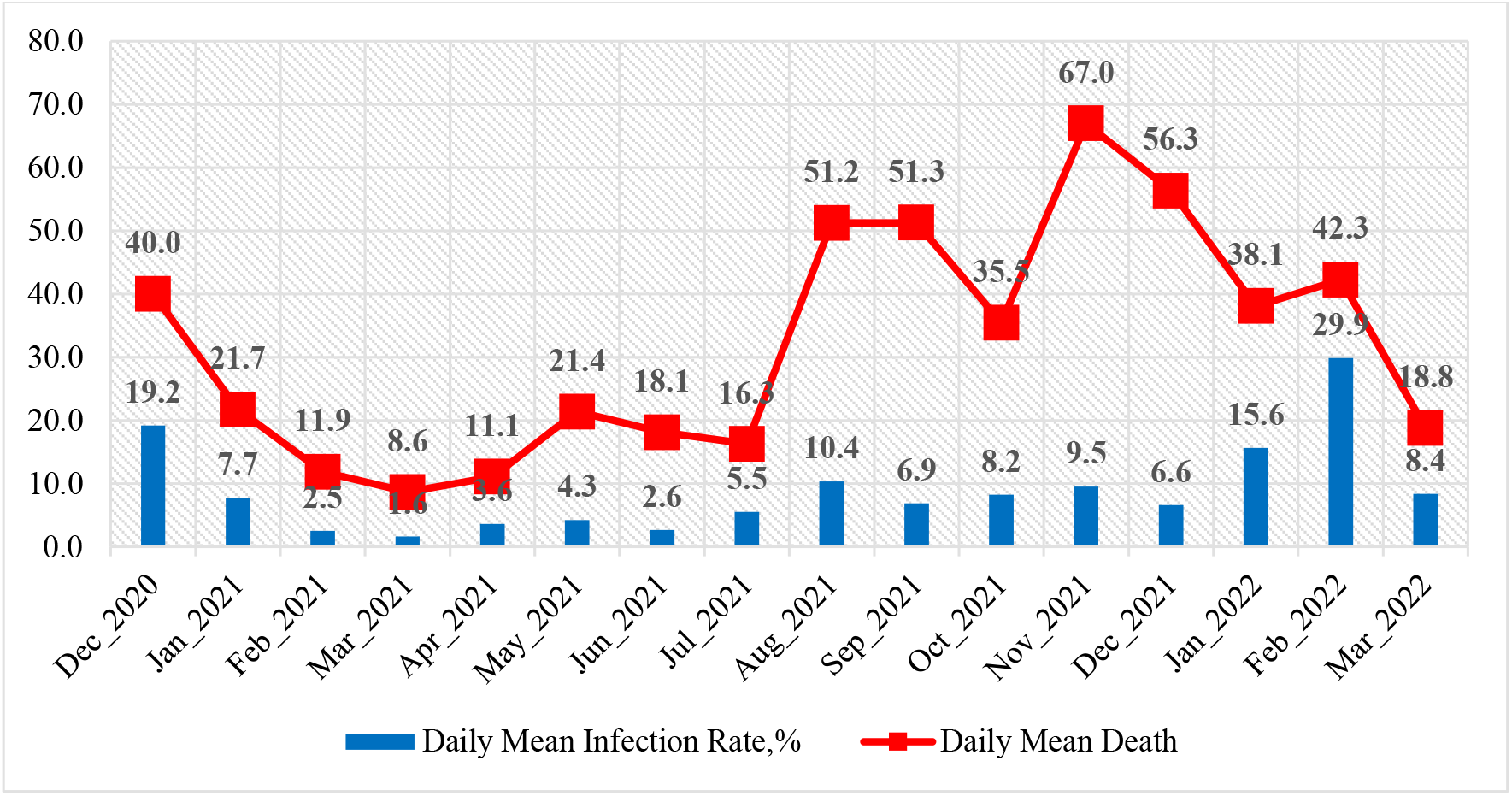
Mean monthly values of COVID-19 infection rate and deaths cases in Georgia from December 2020 to March 2022.

**Fig. 19.**
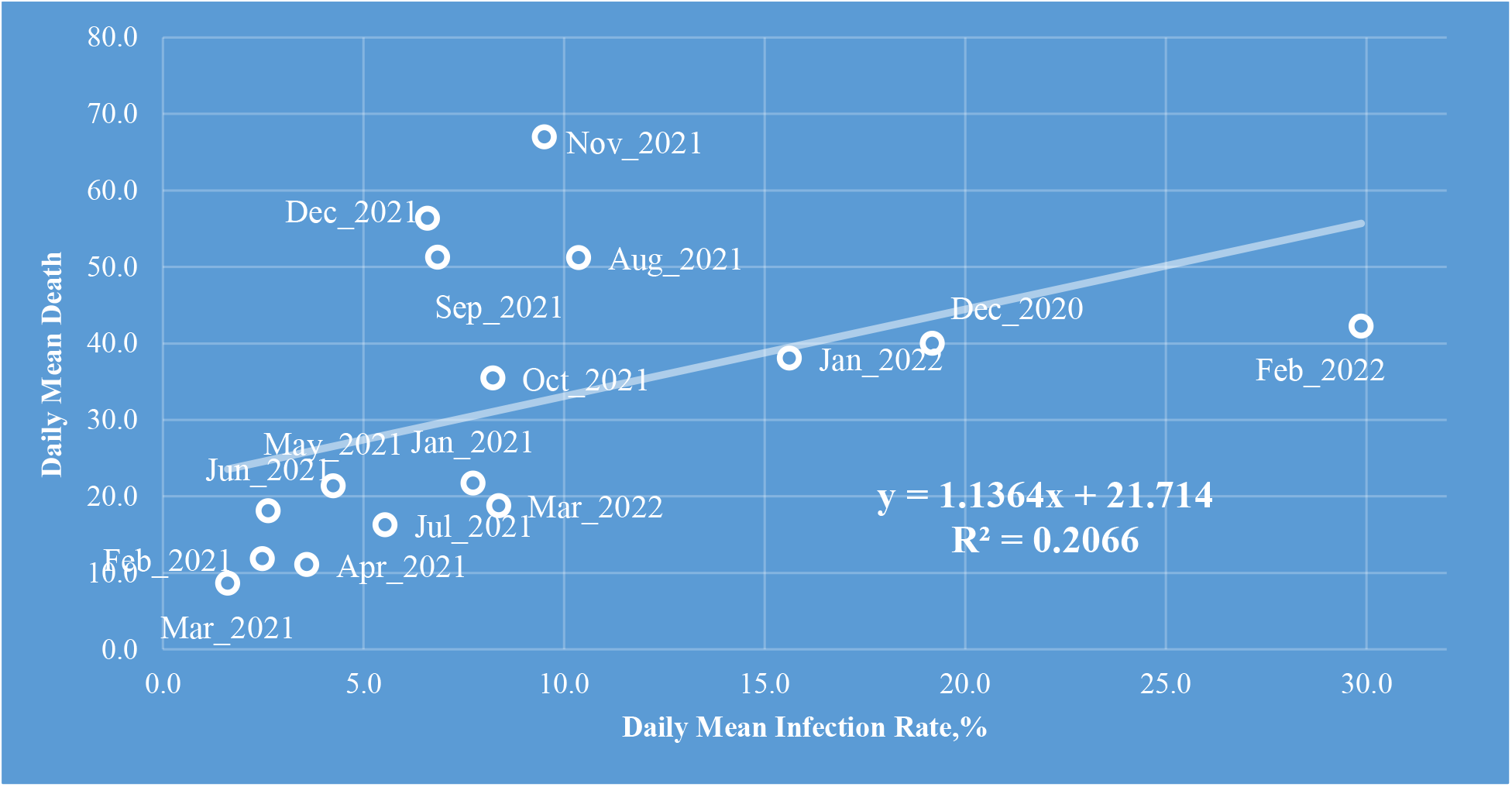
Linear correlation and regression between the mean monthly values of COVID-19 infection rate and deaths cases in Georgia from December 2020 to March 2022.

As noted in [10, 11, 13] and above (Fig. 11 and 12), there is some time-lag in the values of the time series R and D with respect to C. An estimate of the values of this time-lag for January-March 2022 is given below (Fig. 20).

**Fig. 20.**
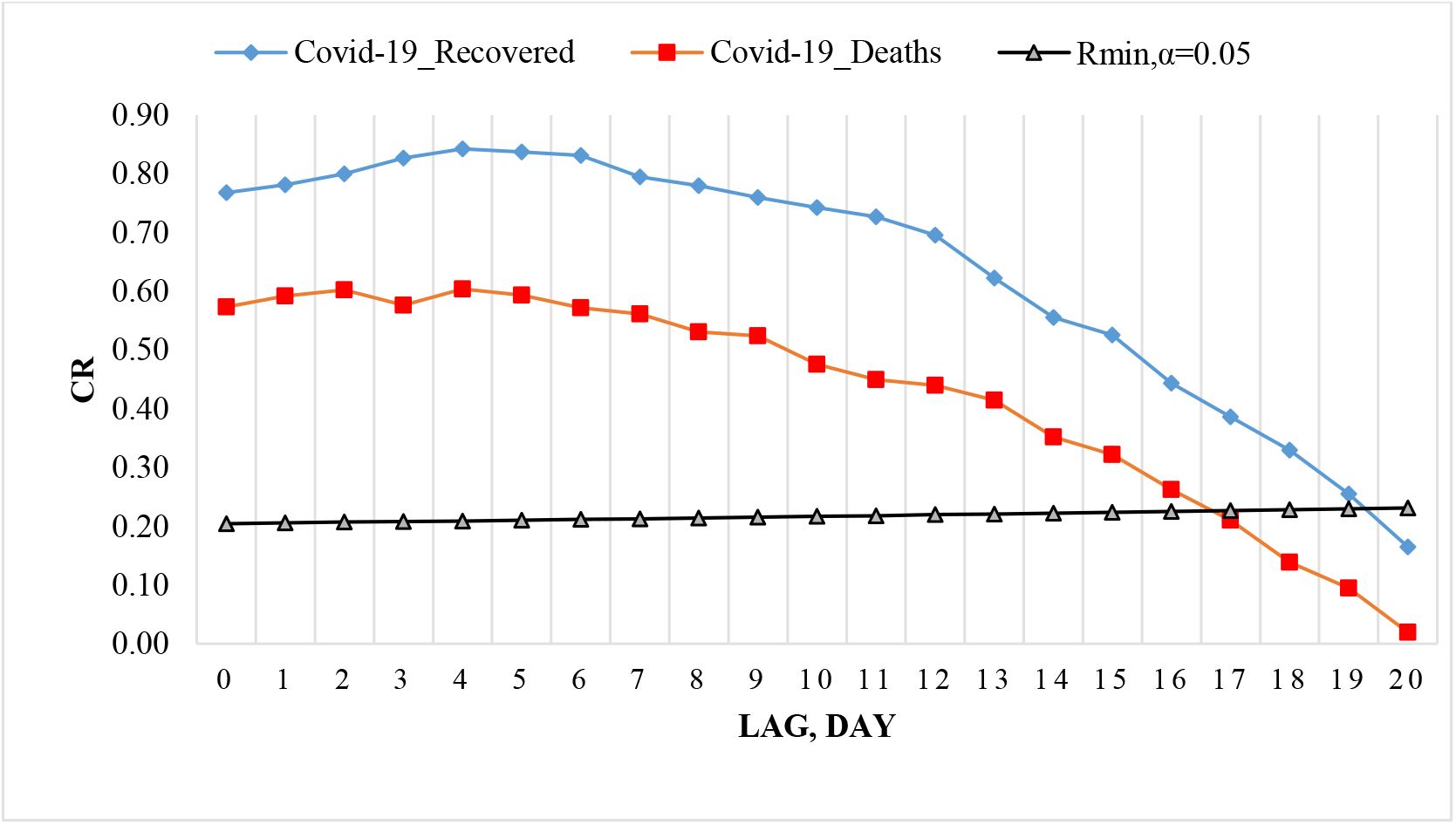
Coefficient of cross-correlations between confirmed COVID-19 cases (normed to tests number) with recovered and deaths cases in Georgia from 01.01.2022 to 31.03.2022.

Cross-correlations analysis between confirmed COVID-19 cases with recovered and deaths cases shows, that in spring 2021 the maximum effect of recovery is observed 9 and 13 days after infection, and deaths - after 12-17 days [10]. In spring 2021 the cross-correlation of the C values with the R and D values weakens completely 38-45 days after infection. Maximum values of CR were 0.81 for R, and 0.79 for D.

In summer 2021 the maximum effect of recovery is observed 19 days after infection (CR =0.95), and deaths - after 16 and 18 days (CR=0.94). In contrast to the spring of 2021, in the summer of 2021, a high level of cross-correlation of C values with R and D values to 40 days after infection is observed (CR ≥ 0.80). In summer 2021 the cross-correlation of the C values with the R and D values weakens completely 56-58 days after infection. So, in Georgia in summer 2021, the duration of the impact of the delta variant of the coronavirus on people (recovery, mortality) could be up to two months [11].

From September 1, 2021 to November 30, 2021 the maximum effect of recovery is observed on 12 and 14 days after infection (CR =0.77 and 0.78 respectively), and deaths - after 7, 9, 11, 13 and 14 days (0.70≤ CR ≤0.72); from October 1, 2021 to December 31, 2021 - the maximum effect of recovery is observed on 14 days after infection (CR =0.71), and deaths - after 9 days (RC=0.43). In Georgia from September 1, 2021 to November 30, 2021 the duration of the impact of the delta variant of the coronavirus on people (recovery, mortality) could be up to 28 and 35 days respectively; from October 1, 2021 to December 31, 2021 - up to 21 and 29 days respectively [13].

From January 1, 2022 to March 31, 2022 the maximum effect of recovery is observed on 3-6 days after infection (CR=0.83-0.84), and deaths - after 2 and 4 days (CR=0.60). The impact of the omicron variant of the coronavirus on people (recovery, mortality) could be up to 19 and 16 days respectively (Fig. 20).

### 3.4 Comparison of real and calculated prognostic daily and monthly data related to the New Coronavirus COVID-19 pandemic in Georgia from January 1, 2022 to March 31, 2022

With September 1, 2021, we started monthly forecasting values of C, D and I. In Fig. 21–24 and Table 8 examples of comparison of real and calculated prognostic daily and mean monthly data related to the COVID-19 coronavirus pandemic in Georgia from January 1, 2022 to March 31, 2022 are presented. Note that as earlier the results of the analysis of monthly forecasting of the values of C, D and I, information about which was regularly sent to the National Center for Disease Control & Public Health of Georgia and posted on the Facebook page https://www.facebook.com/Avtandil1948/.

**Table 8.**
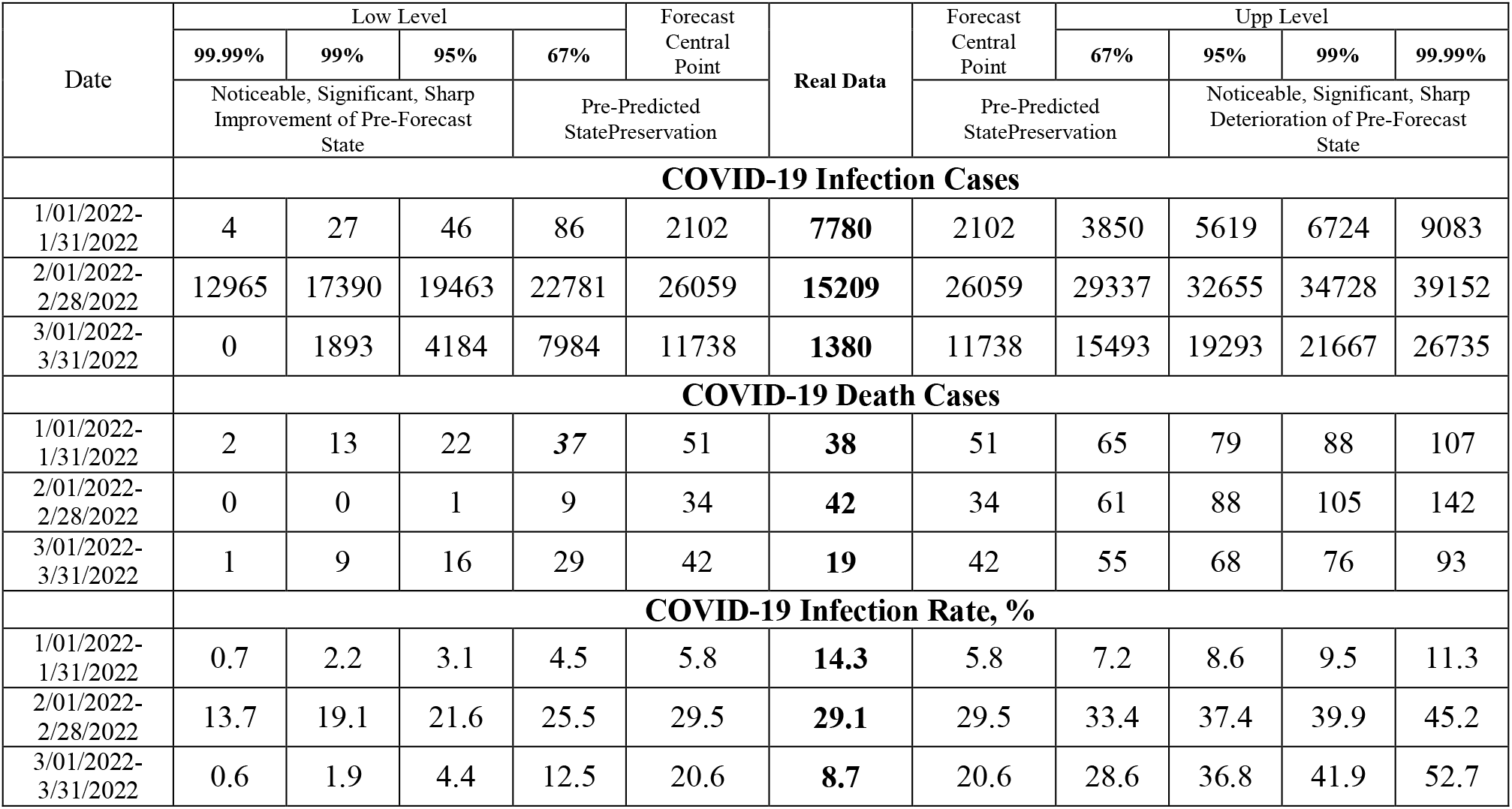
Verification of monthly interval prediction of COVID-19 confirmed infection cases, deaths cases and infection rate in Georgia from 01.01.2022 to 31.03.2022.

**Fig. 21.**
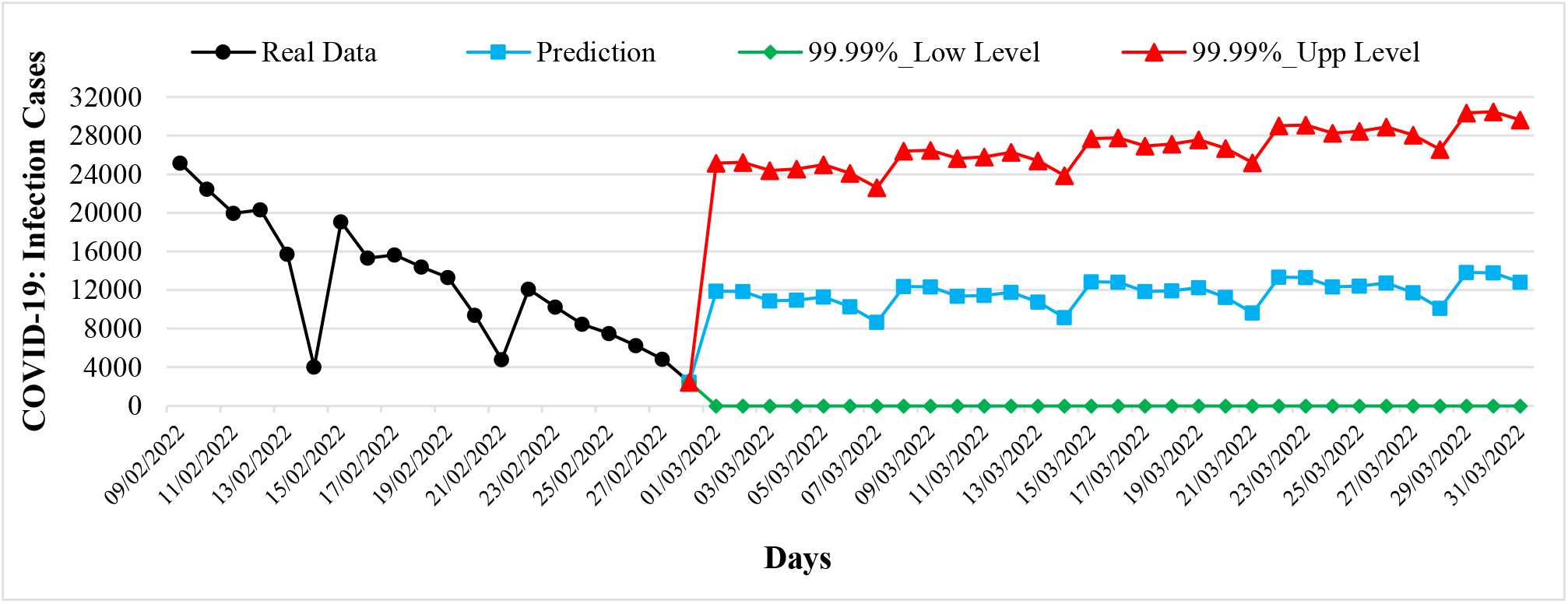
Example of Interval Prediction of COVID-19 Infection Cases in Georgia from 01.01.2022 to 31.01.2022.

**Fig. 22.**
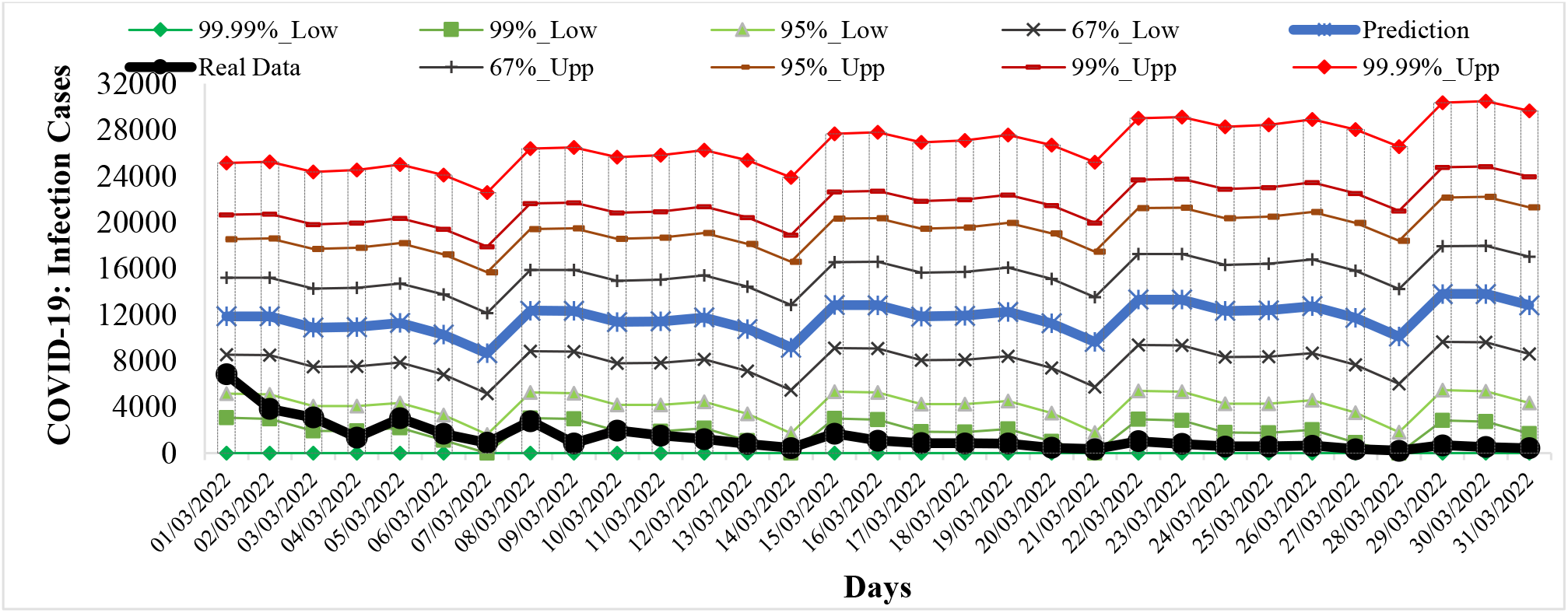
Example of Verification of Interval Prediction of COVID-19 Infection Cases in Georgia from 01.01.2022 to 31.01.2022.

**Fig. 23.**
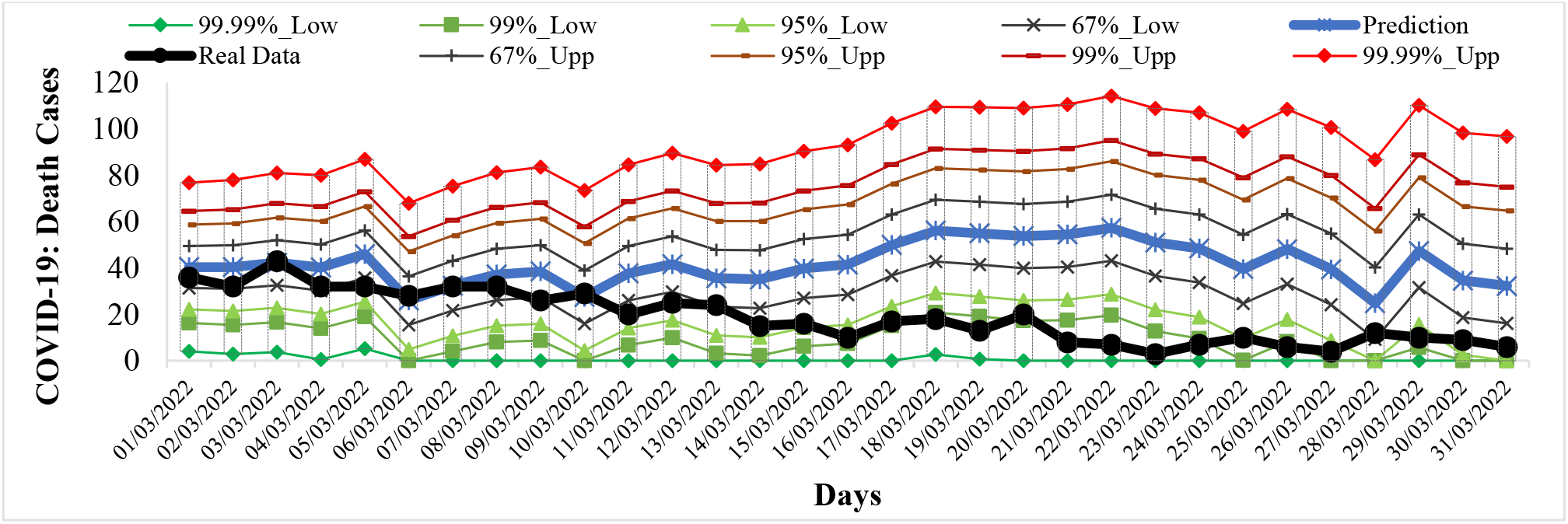
Example of Verification Interval Prediction of COVID-19 Death Cases in Georgia from 01.01.2022 to 31.01.2022.

Comparison of real and calculated predictions data of C, D and I (Fig. 21–24, data from https://www.facebook.com/Avtandil1948/, Table 8) shown, that in investigation period of time daily and mean monthly real values of C, D and I mainly fall into the 67% - 99.99% confidence interval of these predicted values. Exception - predicted values of I for January 2022 (alarming deterioration, violation of the stability of a time-series of observations)

**Fig. 24.**
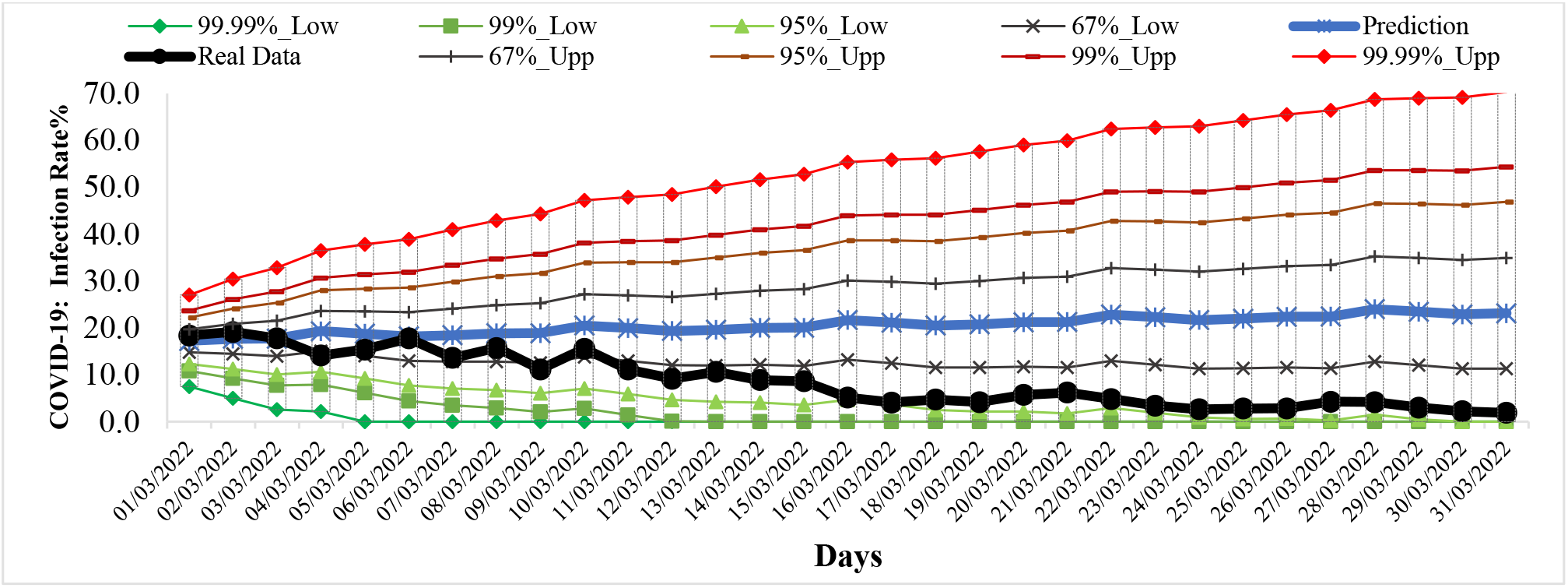
Example of Verification Interval Prediction of COVID-19 Infection Rate in Georgia from 01.01.2022 to 31.01.2022.

For all forecast periods (3 monthly forecasting cases), exception predicted values of I for January 2022 (violation of the stability of a time-series of observations), the stability of real time series of observations of these parameters (period for calculating the forecast + forecast period) remained (Table 9).

**Table 9.** Change in the forecast state of C, D and I in relation to the pre-predicted one according to Table 1 scale [9].

| Date | COVID-19 Infection Cases |
| --- | --- |
| 1/01/2022- 1/31/2022 | Sharp deterioration, stability of a time-series of observations |
| 2/01/2022-2/28/2022 | Sharp improvement, stability of a time-series of observations |
| 3/01/2022-3/31/2022 | Sharp improvement, stability of a time-series of observations |
|  | COVID-19 Death Cases |
| 1/01/2022- 1/31/2022 | Preservation, stability of a time-series of observations |
| 2/01/2022-2/28/2022 | Preservation, stability of a time-series of observations |
| 3/01/2022-3/31/2022 | Noticeable improvement, stability of a time-series of observations |
|  | COVID-19 Infection Rate |
| 1/01/2022- 1/31/2022 | Alarming deterioration, violation of the stability of a time-series of observations |
| 2/01/2022-2/28/2022 | Preservation, stability of a time-series of observations |
| 3/01/2022-3/31/2022 | Noticeable improvement, stability of a time-series of observations |

Thus, as earlier, the daily monthly and mean monthly forecasted values of C, D and I quite adequately describe the temporal changes in their real values.

## Conclusion

In the future until the end of the pandemic it is planned to continue regular similar studies for Georgia in comparison with neighboring and other countries.

## Data Availability

https://www.soothsawyer.com/john-hopkins-time-series-data-with-us-state-and-county-city-detail-historical/; https://data.humdata.org/dataset/total-covid-19-tests-performed-by-country; https://stopcov.ge

https://www.facebook.com/Avtandil1948/

https://stopcov.ge

